# Impacts of green space interventions in educational settings on children and young people’s mental wellbeing: a systematic review

**DOI:** 10.1101/2025.01.09.25320287

**Authors:** Judith Eling, Steven Cummins

## Abstract

In the context of a recent decline in mental wellbeing in children and young people, there is increasing policy and practice interest in the benefits of children’s engagement with the natural environment to improve mental wellbeing. Although there is a growing evidence base linking health outcomes across populations with exposure to nature, the evidence base for the health, wellbeing and educational attainment benefits of green space interventions in children and adolescents is of lower quality. The aim of this systematic review was to assess whether the current evidence enables us to draw conclusions about the relationship between different educational green space intervention types and different categories of mental wellbeing outcome. The review adopted an approach to mental wellbeing grounded in the field of positive psychology, which sees wellbeing as a positive resource, composed of emotional, psychological and social domains. A number of outcomes was grouped in each domain, for example attention and executive function skills were grouped in the psychological wellbeing domain. Green space interventions were categorised into five sub-types: environmental education, break-time or play in a green environment, school gardening, learning in nature and nature pre-school/forest school. Extensive searches of seven databases, updated in March 2024, identified 36 quantitative studies, including quantitative components of mixed-methods studies. Study quality was assessed using the EPHPP tool. Due to study heterogeneity, meta-analysis was not feasible; instead, a Synthesis Without Meta-analysis (SWiM) based on effect direction was performed; additionally, a narrative synthesis summarising the effect direction at study level was conducted where an outcome was only measured within a single study within the same intervention category. Out of 37 reports of 36 studies, two had a strong overall rating, ten had a moderate quality rating and 25 had a weak overall rating. Based on generally very low certainty of evidence, there was some evidence for the positive effects of environmental education and breaks in green spaces on emotional wellbeing outcomes; for beneficial effects of breaks in green spaces on social wellbeing outcomes; and for improved outcomes in particular categories of psychological wellbeing associated with breaks in green spaces, learning in nature and forest school. Weaknesses identified in studies were often linked to specific features in study design, such as intervention dilution, as well as to lack of sensitivity of measurement instruments. This is the first comprehensive systematic review aiming to assess the effects of green space interventions in educational settings on outcomes across all three dimensions of mental wellbeing in children and young people aged 0-18. Recommendations are made on how study design can be improved in order to generate a stronger evidence base, including by having separate control groups, using a larger exposure dose and duration, extending the length of follow-up, and designing more nuanced interventions.

## Introduction

Many high-income countries have poor child wellbeing outcomes, with some of the wealthiest, including the USA, New Zealand and Republic of Ireland in the bottom third of a 41-country wellbeing league table (1). The United Nations Convention on the Rights of the Child states that every child has the right not only to survive, but to live to their fullest potential, developing healthily in conditions that do not adversely affect their physical and mental wellbeing (2). Children’s wellbeing is thought to be critical to a child’s social and educational attainment throughout their school career (3, 4) as well as having important consequences for mental health and social outcomes in adult life (5). However, in many countries children’s mental wellbeing has declined, particularly over the past 4 years, as a result of prolonged collective stress during the COVID-19 pandemic (6) which has disproportionately affected poorer and more vulnerable children (7, 8).

There is increasing policy and practice interest in the benefits of children’s engagement with the natural environment to improve mental wellbeing (9-16). At the same time human populations globally are less likely to have contact with nature due to rapid urbanisation (17), habitat loss and diminishing biodiversity (18), as well as increasing restrictions on children’s independent movements (19). These changes have been felt most acutely by some disadvantaged populations, and inequitable access to green space has been well documented (20-22). Provision of opportunities for children to have contact with nature within educational settings may therefore help address inequalities in access to, and engagement with, nature to improve their health and wellbeing. However, to do this effectively it is important to ascertain which green space interventions in educational settings may be most likely to achieve positive impacts on emotional wellbeing.

Although there have been high-quality evidence syntheses linking health outcomes across populations with exposure to nature (23, 24), the evidence base for the health, wellbeing and educational attainment benefits of green space interventions in children and adolescents is of lower quality (12, 15, 16, 25-34). Some reviews have focused on specific interventions, but with a broad and heterogenous range of outcomes. For example, a mixed-methods review of impacts of school gardening (29) found qualitative studies were of better quality than quantitative, ascribed a range of health and wellbeing benefits to school gardens, and suggested that students who were academically weaker particularly benefited. A review of the mental, physical and social health benefits of immersive nature-experience for children and adolescents (12) found inconclusive results for mood improvement, with conditional support for other outcomes such as resilience and academic performance. However, evidence was of mixed quality with risk of bias, insufficient sampling methods and unsuited comparison groups as common study limitations. A review of the impacts of unstructured nature play on health in early childhood (35) identified only two studies reporting emotional outcomes.

Other reviews have focused on mental wellbeing in association with a broad range of nature interventions. A review of the effects of nature activities on wellbeing in children and young people (36) found that though a range of nature activities may impact on wellbeing outcomes, including positive affect, stress reduction and restoration, results were inconsistent. Some studies did not identify positive changes in certain outcomes, and two out of 14 studies reported negative effects. A recent review of the effect of nature on children’s psychological wellbeing (37) identified predominantly observational studies reporting on the effect of nature exposure in children’s wider environment (residential, as well as school) on wellbeing.

Previous reviews have used thematic findings from qualitative studies to comment on possible causal pathways by which engagement with nature promotes wellbeing (29, 38). A number of potential pathways had been proposed since the early 1980s. In 1981, Ulrich’s *stress reduction theory* (SRT) (39) postulated that natural environments promote recovery from stress, while urban environments tend to hinder the same process. The *biophilia hypothesis* (40), published in 1984, suggested that humans have an innate need to affiliate with nature and that this need has become biologically coded. In the late 1980s Stephen and Rachel Kaplan (41, 42) published their *attention restoration theory* (ART) which suggests that mental fatigue and concentration can be improved by time spent in, or looking at nature. Kuo (43) suggested in 2015 that *enhanced immune functioning* may provide a satisfactory central pathway for nature’s positive impact. Further mechanisms proposed include beneficial effects of exposure to *phytoncides* (naturally occurring chemical compounds secreted by plants) (44), fractals (self-similar patterns occurring in nature) (45), sunlight (46), nature sounds (47), and soil bacteria (48). Building on the biophilia hypothesis, the construct of a measurable *nature connectedness* has been researched more extensively (49), and in particular, the benefits of nature connectedness as a mediator between nature exposure and wellbeing in children (50, 51).Little agreement was found between reviews on what was meant by mental wellbeing, with some identifying a broad range of outcomes across all wellbeing domains, and others focusing on a subset such as emotions or attention restoration. Therefore, a brief definition of mental wellbeing follows for the purposes of this review.

Concepts of mental wellbeing are multifaceted, going back historically to Aristotle’s notion of *eudaimonia* or happiness, to be accomplished by people’s full and balanced realisation of their capabilities (52). More recently, there has been debate about the *eudaimonic* vs *hedonic* dichotomy of discourse on wellbeing (53, 54), with consensus now reached that these are two sides of the same concept (55). In the discourses of the *capabilities approach* to children’s mental wellbeing (56) and of *positive psychology* (57), wellbeing is seen as a positive resource and a separate dimension from mental health (58). The UK’s National Institute for Clinical Excellence (NICE) defines mental wellbeing as falling across three domains: emotional, psychological and social (46).

This review adopts the latter view of wellbeing as being a positive resource, composed of emotional, psychological and social domains. It adds to previous reviews by focusing on green space interventions in educational settings (from ages 0-18) and reporting on the whole range of mental wellbeing outcome domains. It attempts to answer the following questions: 1. What are the impacts of greenspace interventions in educational settings on the mental wellbeing of children and young people under 18? 2. Do these impacts differ depending on the intervention type, duration of intervention and population attributes (such as age, gender, ethnicity and socioeconomic status)? 3. What is the quality of the evidence base, and how does this inform recommendations for future research?

In face of inequalities in access to green space between socioeconomic groups in the wider environment (59), educational settings have the potential to act as levellers, reducing inequalities in health and wellbeing outcomes for children and young people (60). The findings of this review will therefore be useful to inform the design of future research as well as guiding policy makers and practitioners in education and health.

## Methods

This review protocol was prospectively registered with PROSPERO (CRD42020225124), initially as a mixed methods rapid review. An amendment was subsequently accepted on to convert the study to a systematic review of quantitative studies. The findings were reported following PRISMA guidelines (suppl. file B).

### Search strategy

Seven electronic databases were searched: Medline via Ovid, PsycINFO via EBSCOHost, GreenFILE via EBSCOHost, Web of Science, EMBASE, ERIC and Global Health via EBSCOHost. The search strategy was peer reviewed under PRESS guidance (61) and a sample search strategy is attached in suppl. file (B). Databases were searched from inception through to March 6^th^ 2024. The search concepts “green space” (intervention), “educational setting” (population) and “mental wellbeing” were examined using a combination of free-text terms and Medical Subject Heading (MeSH) terms, informed by previous systematic reviews. A grey literature search was conducted using thesis repositories (OATD, Global ETD Search and ProQuest), the English National Grey Literature Collection, the OpenGrey collection (archived in 2021), the online British Library catalogue and on institutional research repositories related to the field of study. A supplementary search was carried out by backward and forward citation chasing, searching key journals, authors’ publication lists and key literature reviews.

### Selection criteria

Both quantitative and qualitative studies were included, if they met the criteria in table 1. Articles were de-duplicated and initially included based on title screening in Endnote9 software. Abstracts were then imported into Rayyan software and assessed against inclusion and exclusion criteria. Screening of titles and abstracts was conducted by one author (JE). Full texts were appraised for any studies where there was any uncertainty regarding inclusion, and these studies were independently reviewed by the second author (SC), with discussion taking place afterwards and consensus being reached where assessments differed. The main reasons for excluding studies at the full text screening stage were: (i) studies not meeting the outcome criteria (for example, where title or abstract had included mental wellbeing, but the actual measures used were physiological wellbeing measures) or (ii) the study design being cross-sectional rather than longitudinal or (iii) studies not meeting the intervention criteria (for example, where the intervention had a residential component).

**Table 1:** PICO inclusion and exclusion criteria.

|  | **Inclusion criteria** | **Exclusion criteria** |
| --- | --- | --- |
| **Population** | Child and adolescent populations under the age of 18<br>Studies reporting on populations under as well as over the age of 18 were included only if they reported the age groups separately, and if the majority of participants were under 18. | Population aged 18 and over |
|  | Studies investigating impacts of interventions on particular groups which were representative of their local geography (such as children from a low socio-economic background) were included | Studies investigating impacts of interventions on particular subgroups such as children with ASD or ADHD were excluded. |
| **Intervention** | Studies were included if the intervention reported on was taking place in an educational setting or organised by an educational institution. |  |
|  | After-school clubs held in educational settings, such as gardening clubs, were included as felt to be accessible and equitable. | Residential interventions were not included as they were not deemed equitably accessible, as well as being potentially susceptible to significant co-intervention bias. |
|  | The following intervention typologies were included:<br>• Break/playtime in a green environment, including playground greening interventions<br>• School gardening<br>• Nature education, defined as any teaching activity whose aim was to promote children's exploration of, and to increase the understanding of, nature.<br>• Environmental education activities (activities explicitly aiming to increase students' understanding of environmental | Indoor nature interventions such as green walls, or views from the school building onto nature |
|  | issues). Such studies were included if the activities took place outside the school site, if they were part of the school timetable.<br>• Learning in nature, defined as the delivery of the standard school curriculum outside the classroom, in a natural setting.<br>• Forest school or nature preschool, defined as accomplishing child development goals through experiences in, and with nature in a woodland or natural setting |  |
| **Comparators** | Non-participation in an activity in a green space taking place in an educational setting. |  |
| **Outcomes** | Mental wellbeing outcomes (encompassing emotional, psychological and social domains) measured either using a uni-dimensional tool or scale, or as a component of a global wellbeing or mental wellbeing scale (studies where results of global scales were not disaggregated have been included, but results have been annotated). | Academic achievement outcomes |
|  | Studies viewing mental wellbeing as a positive asset were included | Studies measuring absence of mental illness diagnoses, such as anxiety and depression, were not included |
|  |  | Studies reporting any purely physiological outcomes, such as physiological measures of stress, were excluded. |
| **Other** | Quantitative studies were included if the study design was a randomised controlled trial, non-randomised controlled trial, and any other before-and-after study. | Studies with a cross-sectional design |
|  | Studies published in English | Studies published in languages other than English |
|  | Studies conducted in Europe, North America, Australia and New Zealand | Studies conducted in any other country |
|  |  | Studies with no formal testing of differences |

### Data extraction and quality appraisal

Data items were extracted from the literature by a single reviewer (JE) using a pre-piloted data extraction template (appendix). The categories recorded were type of source, study design, country, inclusion and exclusion criteria, recruitment, sample size and characteristics, setting, description of the intervention and duration, comparator, outcome measures, main findings, study limitations and sources of bias. Any unclear results were discussed with the co-author (SC) and consensus reached. The summary of the extracted data can be found in suppl. file D (summary) and suppl. file E (results).

The Effective Public Health Practice Project (EPHPP) tool (62) was used to assess the quality of included quantitative studies.

### Data synthesis

It was not possible to perform a meta-analysis due to heterogeneity and studies not providing the appropriate data to support standardising. Instead, a Synthesis Without Meta-analysis (SWiM) based on effect direction was performed (63). This approach addresses the question of whether there is evidence of a positive or negative association. Findings were presented in a summary table (effect direction plot), with outcomes grouped by similar outcome categories within each outcome domain.

Outcomes were grouped into the three NICE domains of mental wellbeing: (i) emotional (categories: emotional wellbeing, self-esteem, mood/affect/enjoyment and HRQoL/life satisfaction), (ii) psychological (categories: attention/attention restoration/stress recovery, executive function skills (memory/inhibition control/cognitive flexibility) and perceived restorativeness) and (iii) social (categories: perceptions of safety/injuries/bullying/gang activities, positive student interactions/pro-social behaviour/involvement and resilience (initiative/self-regulation/attachment)).

The three effect direction plots present study level effect direction and, where more than one outcome was reported within an outcome category, it presents the overall outcome effect direction, following the methodology utilised in a previous review (38, 64).

In addition, a narrative synthesis summarising the effect direction at a study level was conducted where an outcome was only measured in one study within the same intervention category.

## Results

### Included studies

After duplicates were removed, 52,412 articles remained, of which 51,787 ineligible articles were removed, leaving 625 abstracts to be screened. Of these, 483 abstracts were excluded, leaving 142 full-text articles to be screened. A total of 37 reports, from 36 unique studies, were retained in the final data set (PRISMA flow diagram figure 1).

**Figure 1:**
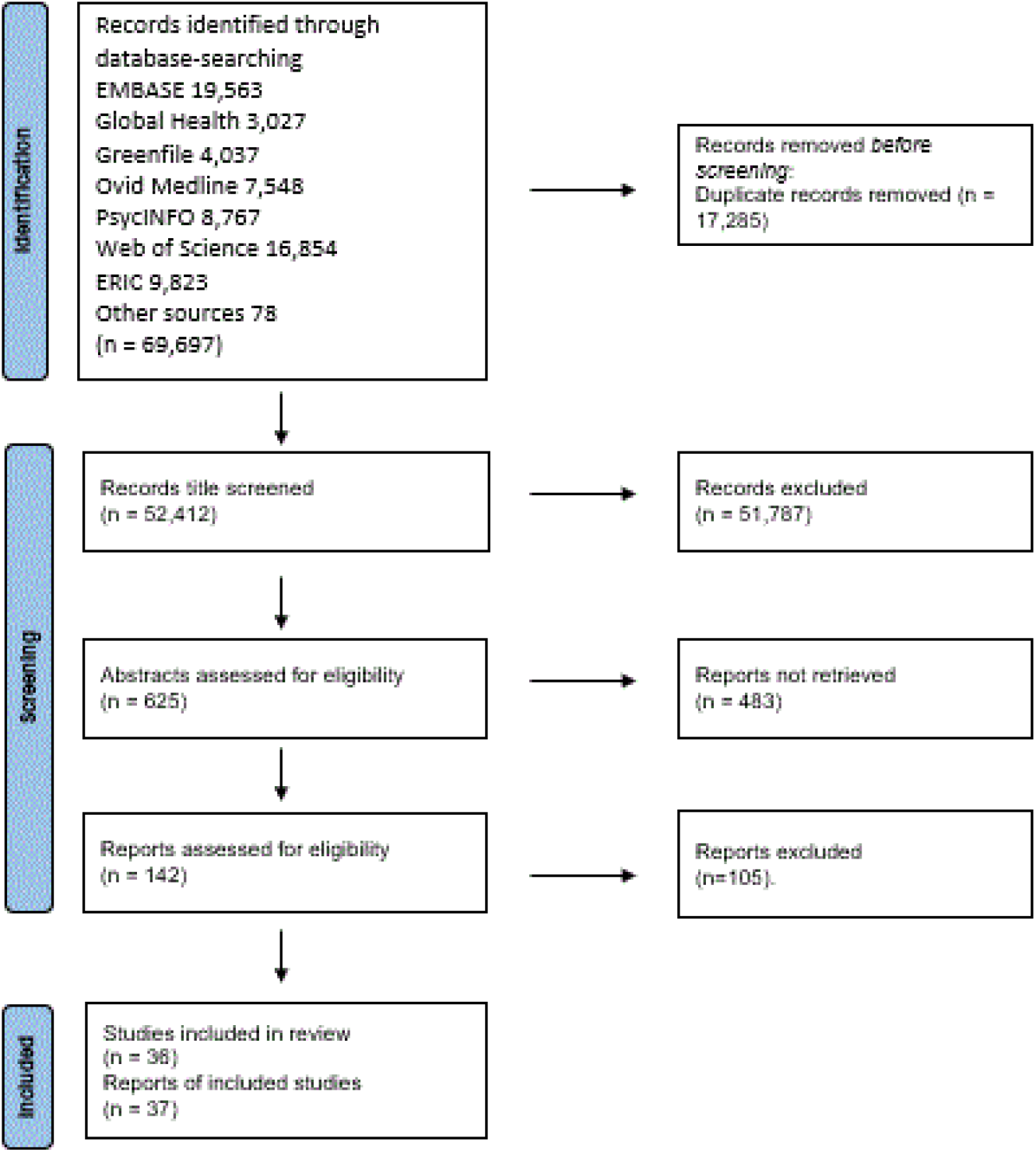
PRISMA flow chart

### Characteristics of included studies

#### Study design

The majority of the included quantitative studies (or quantitative components of mixed methods studies) employed a before-and-after design with a separate control group (n=16, 44%), 13 studies were before-and-after studies with a cross over design (36%)and the remainder employed a before-and-after design without controls (n=7, 20%).

#### Sample size

The total sample size of the combined studies was n=10,295. For controlled studies where this information was provided, intervention arms comprised n=2,064; control arms n=1,014. Many studies either did not have controls or had a crossover design. The majority (n=26, 72%) of studies were small or medium (sample size <300 pupils in the intervention group) with ten studies (28%) having intervention group sample sizes greater than n=300.

#### Participant characteristics

The included studies examined a diverse range of age groups from 0-18. Of the 18 studies where a mean age is reported, the largest group were those reporting a mean age from 5-9 years (n=9; 50%); followed by age 10-14 (n=6; 33.%), age 0-4 (n=2, 11%) and 15-17 (n=1, 6%). Of the 25 studies reporting sex or gender proportions, n=20 (80%) reported the proportion of males or boys as between 48%-54%, n=3 (12%) reported the proportion of males or boys as greater than 55%, n=1 (4%) reported it as less than 45%, and n=1 (4%)reported missing data for sex or gender. Only six studies reported the percentage of students belonging to minority ethnic groups and two studies reported the percentage of students from low socioeconomic groups.

#### Intervention characteristics

Most studies were conducted solely in primary schools (n=16; 44%), followed by studies conducted in early years settings (n=8; 22), studies conducted in both primary and secondary schools (n=7; 20%) and studies conducted solely in secondary schools (n=5; 14%).

The countries where most interventions took place were the USA/Canada (n=13; 36%), the UK (n=10; 28%), Australia/New Zealand (n=4; 11%), Italy (n=4; 11%) with the remaining studies conducted in Spain, Austria, Germany, the Netherlands and Denmark (n=5; 14%).

Intervention types identified by the authors are described in table 2. The most common intervention types were “play or breaks in a natural environment” (n=12; 33%) and “environmental education” (n=9; 25%), followed by “learning in nature” (n=8, 22%) and “forest schools” (n=4; 11%) and “school gardening” (n=3; 9%).

**Table 2:**
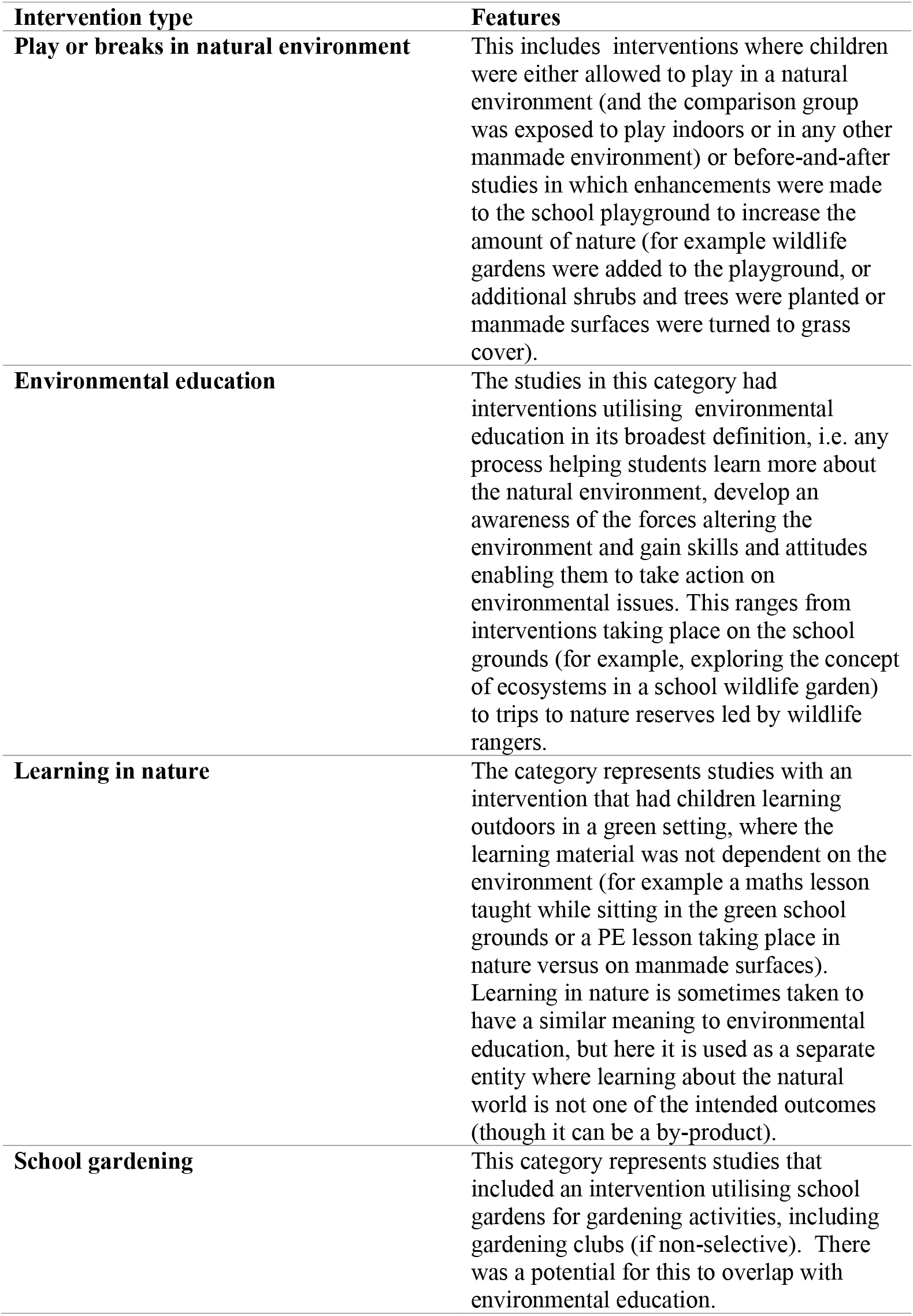
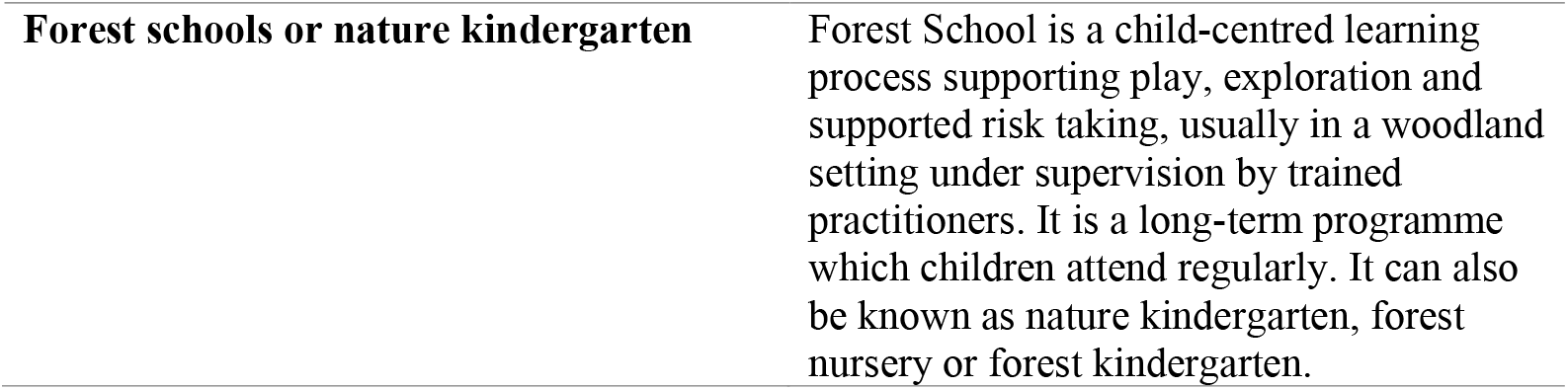
Intervention types.

### Quality of included quantitative studies

The quality of each quantitative study, was assessed by the EPHPP tool (65); details can be found in suppl. file C. Of the 37 reports of 36 eligible studies, two had a strong overall quality rating, ten had a moderate quality rating and 25 had a weak overall rating. Figure 2 presents the quality ratings for all included studies for each EPHPP domain and for the overall rating. Typically, studies were rated weak because of selection bias, issues around confounding, lack of blinding and poor reporting of withdrawals and dropouts. Studies performed relatively well on use of valid and reliable data collection methods.

**Figure 2:**
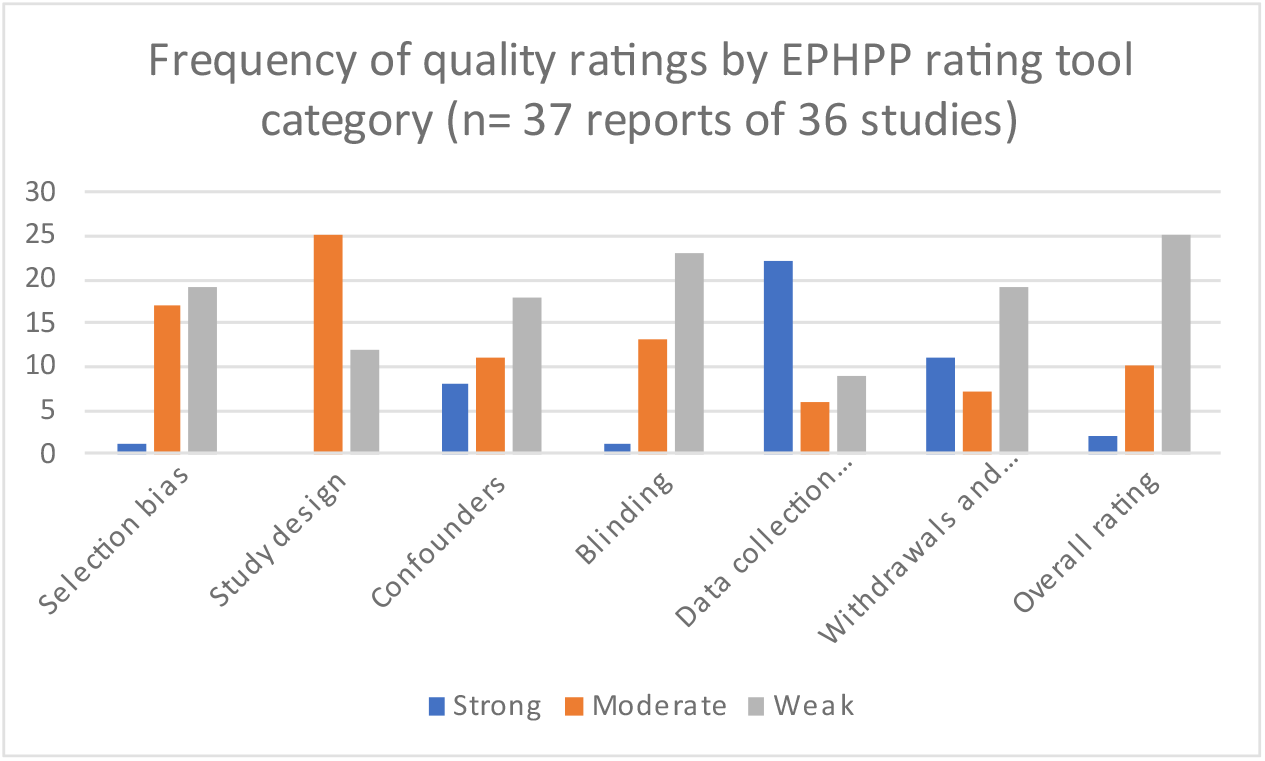
Quality ratings as assessed by EPHPP tool

### Main Findings

Tables 3-5 present the effect direction plots for the three wellbeing domains, per outcome category. Findings with additional details, including effect sizes and confidence intervals, for individual studies can be viewed in suppl. file C.

**Table 3:**
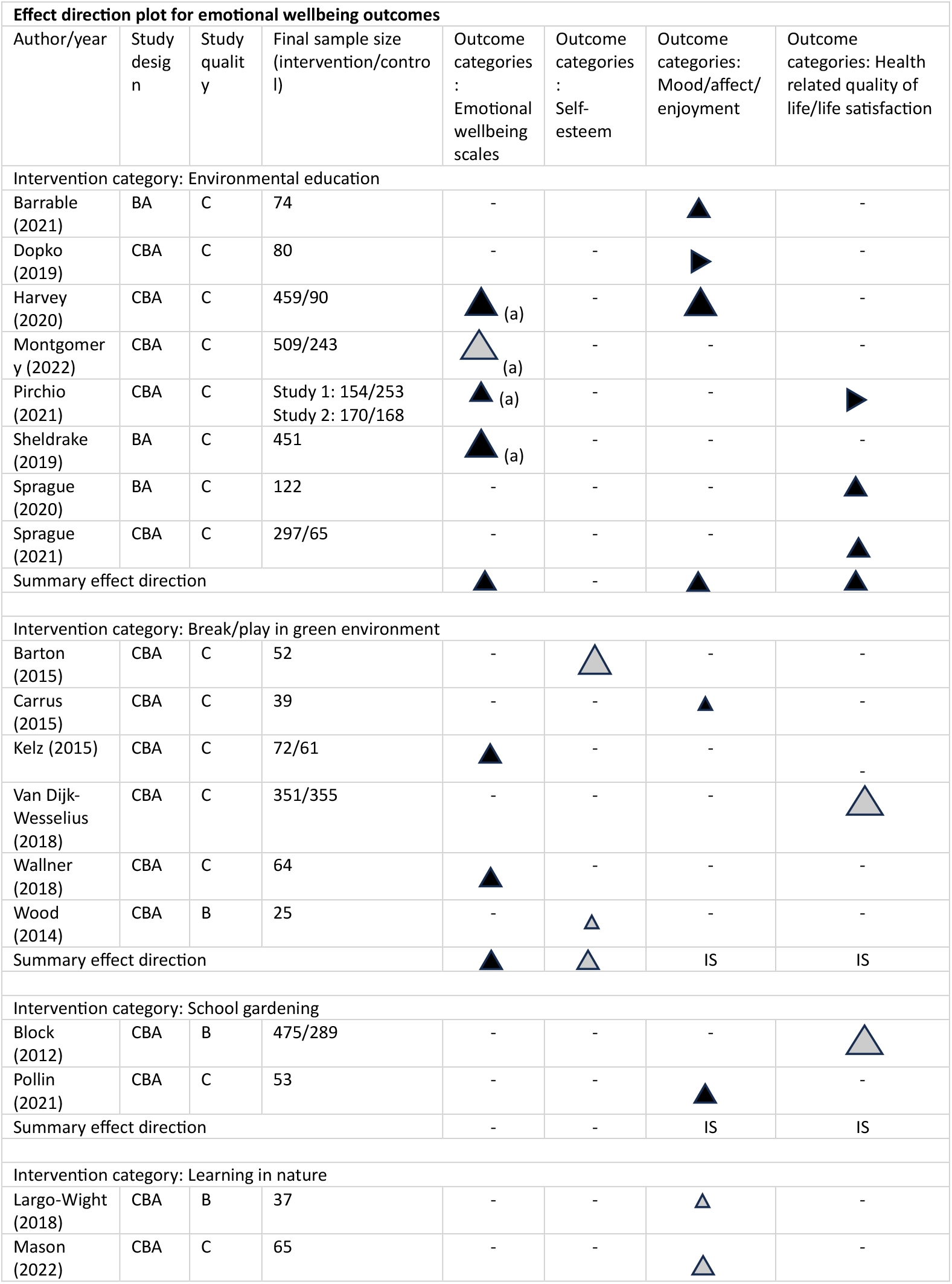
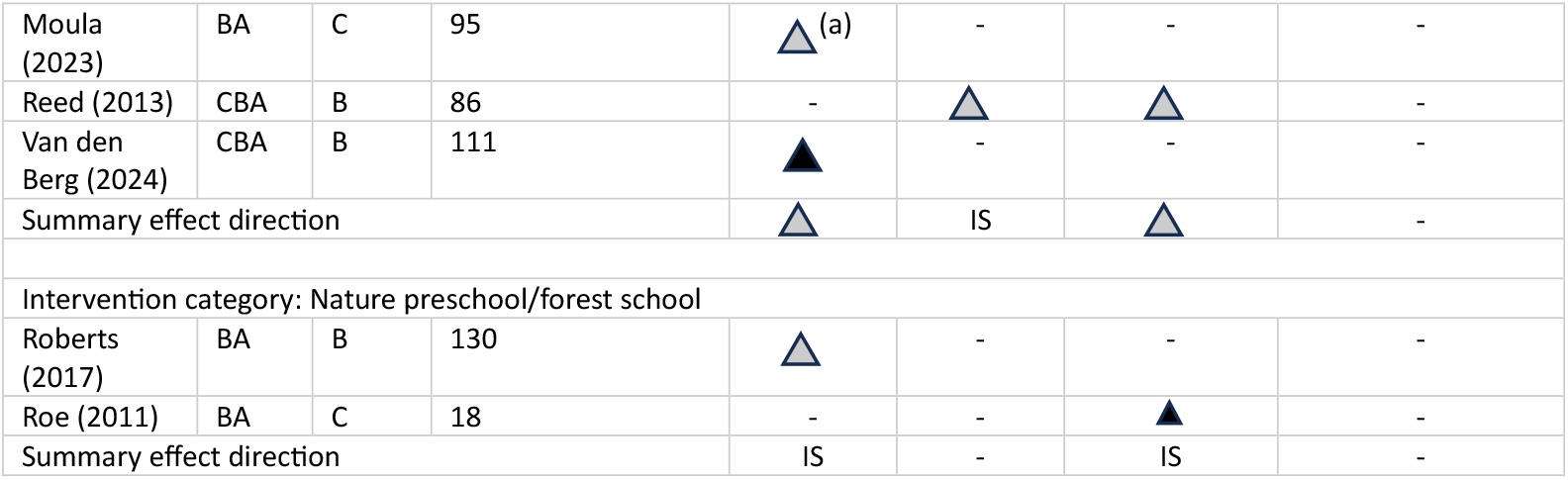
Emotional wellbeing outcomes.

**Table 4:**
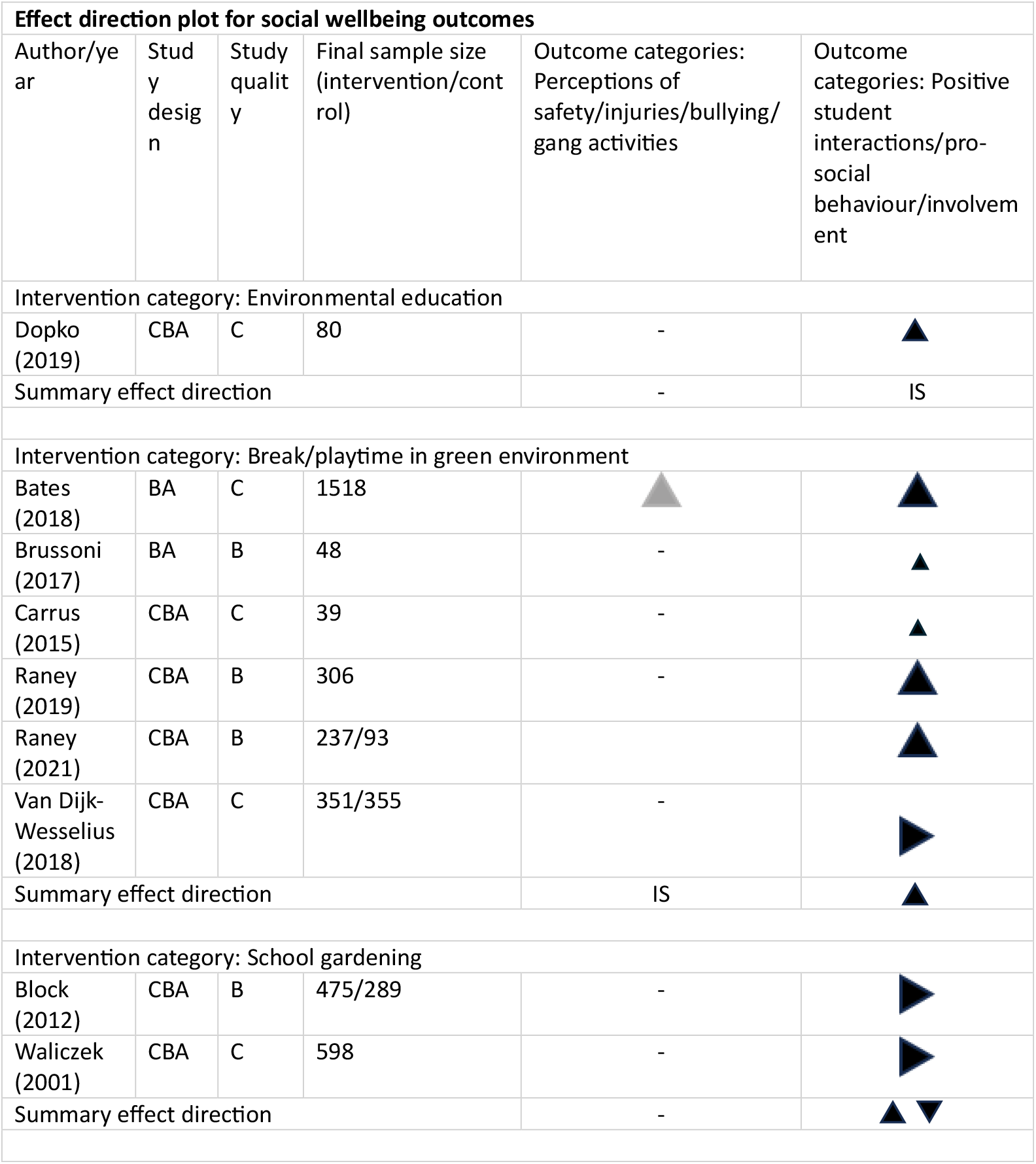
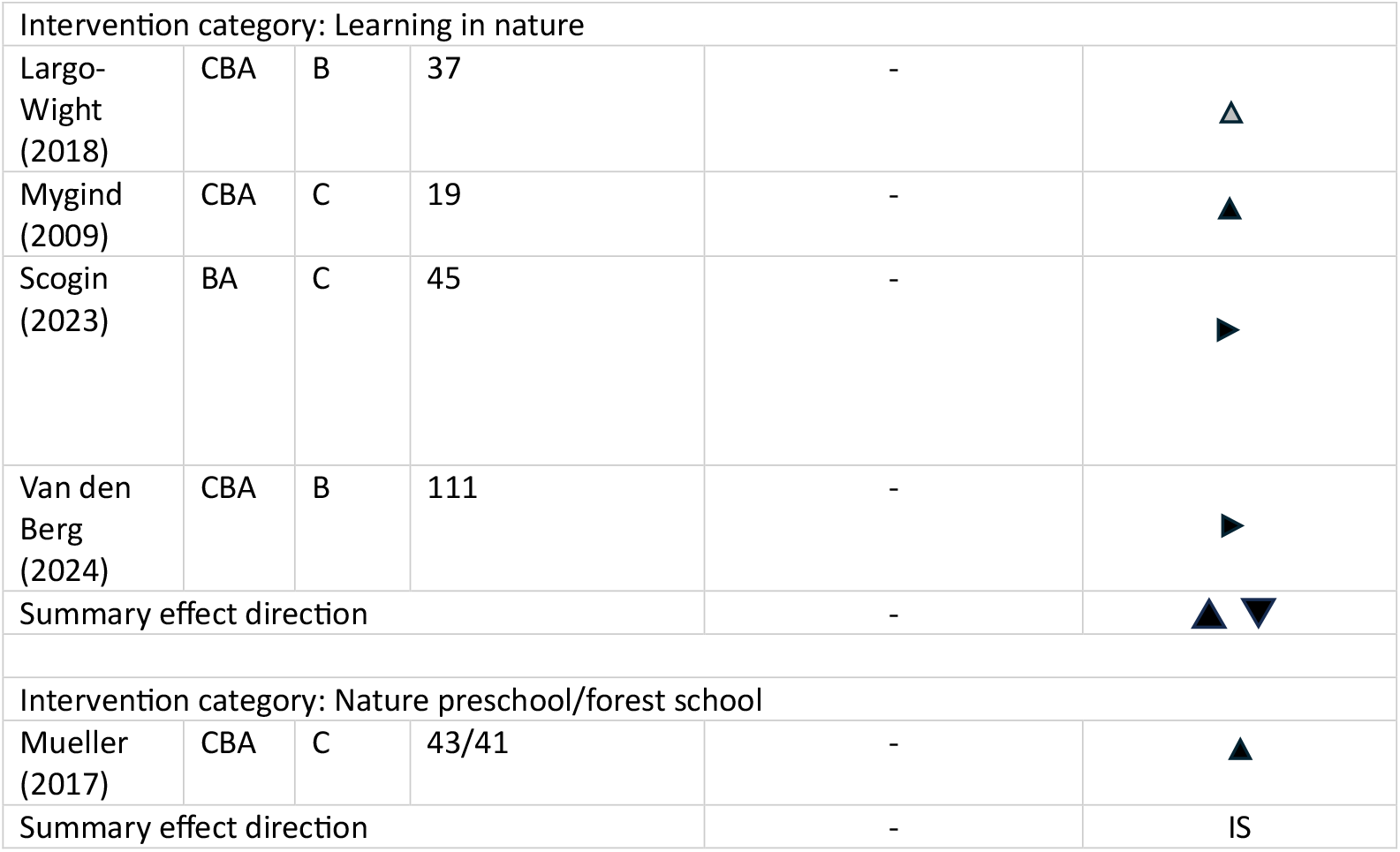
Social wellbeing outcomes.

**Table 5:** Psychological wellbeing outcomes.

| Effect direction plot for psychological wellbeing outcomes |  |  |  |  |  |  |
| --- | --- | --- | --- | --- | --- | --- |
| Author/year | Study design | Study quality | Final sample size (intervention/control) | Outcome: Attention/attention restoration/stress recovery | Outcome: Executive function skills (memory/inhibition control/cognitive flexibility) | Outcome categories: Resilience (initiative/self-regulation/attachment) |
| Intervention category: Environmental education |  |  |  |  |  |  |
| Montgomery (2022) | CBA | C | 509/243 | - | - | ► |
| Summary effect direction |  |  |  | - | - | IS |
| Intervention category: Break/playtime in green environment |  |  |  |  |  |  |
| Amicone (2018) | CBA | B | Study 1:82<br>Study 2: 36 | ▲ | ► | - |
| Anabirtat e (2021) | CBA | B | 167 | ► | - | - |
| Kelz (2015) | CBA | C | 72/61 | ► | - | - |
| Van Dijk-Wesselius (2018) | CBA | C | 351/355 | ▲ | - | - |
| Wallner (2018) | CBA | C | 64 | ▲ | - | - |
| Summary effect direction |  |  |  | ▲ | IS | - |
| Intervention category: School gardening |  |  |  |  |  |  |
| No studies with these outcome categories |  |  |  |  |  |  |
| Intervention category: Learning in nature |  |  |  |  |  |  |
| Largo-Wight (2018) | CBA | B | 37 | ► | - | - |
| Mason (2022) | CBA | C | 65 | ▲ | - | - |
| Scogin (2023) | BA | C | 45 | ▲ | ▲ | ▲ |
| Taylor (2020) | BA | A | 135 | - | ▲ | - |
| Summary effect direction |  |  |  | ▲ | ▲ | IS |
| Intervention category: Nature preschool/forest school |  |  |  |  |  |  |
| Ernst (2019) | CBA | C | 78/14 | - | ► | ▲ |
| Mueller (2017) | CBA | C | 43/41 | ► | ► | - |
| Roberts (2017) | BA | B | 130 | - | - | ▲ |
| Summary effect direction |  |  |  | IS | ▲ ▼ | ▲ |

#### 1) Environmental education

##### a) Emotional wellbeing domain

###### i) Emotional wellbeing scale outcomes (n=4 studies)

Four studies reported a positive effect of environmental education on emotional wellbeing (measured on wellbeing scales); three were significant (66-68) Of these, one (66) reported a significant overall difference in wellbeing scores over time between intervention and control groups for an intervention delivered weekly in school grounds over a 7 week period (p=0.004), however the change in wellbeing scores was statistically significant only in those children who had low wellbeing scores initially. The study with a non-significant finding (69) used a similar design and population to this but reported no significant differences in the changes in the wellbeing scores (p>0.05) between the intervention and control groups. The third study (68), a before-and-after study without controls, consisted of two study cohorts, both of which were reported as having a significant improvement in psychological wellbeing outcomes after four sessions of environmental education in a nature reserve (p=0.000). The fourth study (67) also reported a significant improvement on a wellbeing scale (p<0.001) after visits to wildlife sites, with a significant interaction effect between baseline wellbeing and time.

###### ii) Positive affect (n=3 studies)

Two studies (66, 70)demonstrated a significant positive association between positive affect and environmental education One of these was a before-and-after study without controls (66) which reported a significant increase in positive affect (mean diff: 4.63; CI: 3.49,5.77) and a slight decrease in negative affect (mean diff: +-1.41; CI: -2.09,-0.73). The second study had a controlled before and after design, however only children in the intervention group completed mood surveys, with a significant beneficial effect on positive affect (p < 0.001). A third study (71) reported conflicting results, with both positive (mean diff: 0.28; CI: 0.03-0.54) and negative affect (mean diff: 0.44; CI: 0.18-0.71) increasing significantly in the nature setting versus the control, which the authors hypothesised may have been due to some children experiencing fear and anxiety in the nature setting.

###### iii) Health-related Quality of Life (HRQoL) (n=3 studies)

A controlled before-and-after study (67)did not find a significant effect of contact with nature, resulting from four monthly visits to nature reserves, on life satisfaction. The authors speculated that this might have been due to the timing and intensity of the intervention. Two studies, using similar populations and interventions, used different study designs to investigate the effect of monthly field trips combined with classroom based environmental education on HRQoL domains. A before-and-after study (72) found a significant improvement in emotional health functioning after the intervention (p<0.001). The controlled before-and-after study (73) found a significant improvement in emotional functioning in the intervention group (p<0.001) whereas the control group showed significant reductions in the score, hypothesised by the authors as being due to change in the seasons, from spring to autumn, during the study period.

##### b) Social wellbeing domain

###### i) Positive student interactions/prosocial behaviour/involvement (n=1 study)

In a study exploring primary school children’s pro-social behaviour after a visit to a nature school, children had significantly higher pro-social scores on the majority of tasks, compared with their scores after a visit to an aviation museum (71).

##### c) Psychological wellbeing domain

###### i.) Resilience (n=1 study)

A single, large UK study exploring this outcome did not find a significant change in resilience scores after participation in regular lessons about biodiversity in the school grounds over 21 weeks (69), compared to a control group.

#### 2) Break/play in green environment

##### a) Emotional wellbeing domain

###### i.) Emotional wellbeing scale outcomes (n=2 studies)

One study (74), using a controlled before-and-after design, reported a significant increase in mean wellbeing scores after an intervention to green the schoolyard, and a significant difference in the change between intervention and control groups (p=0.006). A before-and-after study with a crossover design (75) found a significantly reduced reduction in mood lowering in secondary school students who had their break in forest settings compared to students who had their break in a small or medium sized urban park (p=0.001).

###### ii.) Self-esteem (n=2 studies)

Neither of the two eligible studies in this group demonstrated a significant change in self-esteem as a result of play in green spaces. A before-and-after study with a crossover design (76) did not find a significant difference in self-esteem after a nature-based orienteering activity in school grounds, compared to free play on a concrete playground (p>0.05). The second study, using the same design, also did not find a significant change in self-esteem after play in green areas of the school playground, compared to concrete areas (p>0.05).

###### iii.) Positive affect (n=1 study)

A before-and-after study with a crossover design (77) reported a significant increase in positive individual behaviours in preschool children after a free-play activity in nature (p=0.007), which did not occur after free-play in the indoor preschool environment.

###### iv.) HRQoL (n=1 study)

A large (n=2,031children) controlled before-and-after study (78) did not detect a significant change in outcomes on the emotional subscale of the paediatric QoL scale, 48 months after greening the playgrounds of a number of schools (p>0.05).

##### b) Social wellbeing domain

###### i.) Perceptions of safety/injuries/bullying/gang activities (n= 1 study_

A single study (79) reported outcomes on negative social behaviour, and did not find any significant changes in a number of measures (gang-related activity, bullying, injuries) reported retrospectively by teachers, after renovation and greening of primary and middle school playgrounds.

###### ii.) Positive student interactions/pro-social behaviour/involvement (n=6 studies)

Six studies (77-82) reported results on these outcomes, and overall, a positive and significant association was found between children playing in a green environment, and positive social behaviour. Two large studies (81, 82) found a significant decrease in antisocial behaviour and increase in prosocial behaviour 4 months and 16 months after a primary school playground greening measure, and a third large study (79) found that pro-social interactions had increased significantly 6 months after greening and renovation of playgrounds. Two small studies in early years settings found a significant increase in positive relational behaviour when children were able to engage in free play in nature areas (77), and a significant increase in prosocial behaviour after green modification of play areas (80). However, a large study of the effects of playground greening in the Netherlands (78) did not find a significant impact of greening on pro-social orientation, nor on the majority of aspects of social functioning. The authors conceded that greening of schoolyards was modest in some cases and may have led to an underestimation of the effect of greening.

##### c) Psychological wellbeing domain

###### i.) Attention/attention restoration (n=5 studies)

Five studies (74, 75, 78, 83, 84) in this category reported these outcomes, with an overall positive significant effect associated with playing in green spaces. Two studies reported significant increases in attention measures after play, breaks or games in green spaces compared to a control condition (75, 83), whilst a third study found that attention restoration had increased significantly at the study’s second follow-up point, 2 years after greening (78). However, two studies did not find a significant effect: a medium sized study in Spain did not find any evidence that participation in a relaxing activity in a green versus a grey space affected attention restoration (84), and a middle school study in Austria (74) found a small, non-significant (p=0.053) improvement in recovery from stress in a greened secondary school playgrounds after renovation.

#### 3) School gardening

##### a) Emotional wellbeing domain

###### i.) Positive affect (n=1 study)

A study which taught the standard school science curriculum in the course of hands-on gardening lessons and compared this intervention with the same lessons taught in a classroom (85), reported that three out of ten emotions (happiness, pride, surprise) were measured significantly more often after the school garden lessons (p<0.001), and two emotions (disgust, anxiety) were measured more often in the classroom (p<0.05).

###### ii.) HRQoL (n=1 study)

A large (n=475 children) controlled before-and-after study (86) reported a statistically non-significant difference in mean quality-of-life scores between the intervention group, who had participated in a weekly school kitchen garden programme, and the control group (mean diff: 1.23; CI: -0.2 to 2.7; p = 0.09).

##### b) Social wellbeing outcomes

###### i.) Positive student interactions/pro-social behaviour/involvement (n=2 studies)

Two large controlled before-and-after studies (86, 87) examined the impact of school gardening on positive social interactions, and both found no difference in these outcomes between children who participated in a school kitchen garden programme and those who did not. The authors of one of the studies (86)commented that there may have been a “ceiling effect”, with high child cooperative behaviour scores recorded at baseline.

#### 4) Learning in nature

##### a) Emotional wellbeing outcomes

###### i.) Emotional wellbeing scales (n=2 studies)_

Of the two studies examining the effects of learning in nature on outcomes on emotional wellbeing scales, one study (88) reported a significant effect on wellbeing associated with instructed activities in an outdoor nature area in a preschool daycare centre, compared with play outdoors without caregiver interaction; the other study (89) found a small, non-significant effect (p=0.28) associated with an art-in-nature intervention in a primary school. The summary effect direction was therefore a positive association with the intervention, but effects were not statistically significant.

###### ii.) Self-esteem (n=1 study)

A before-and-after study with a crossover design (90) did not find a significant effect on self-esteem of a physical exercise lesson in nature versus in the school grounds (p = 0.72).

###### iii.) Positive affect (n=3 studies)

None of the three studies examining the impact of learning in nature on mood found a significant effect. A small (n=37 children) before-and-after study with a crossover design (91) did not find a significant difference in outcomes between children being taught in outdoors nature versus being taught in an indoor classroom (t = 1.71, p = 0.097). The authors hypothesised that participant fatigue relating to the measurement tool might have been a contributing factor. The second study, using the same study design, found no significant difference in change in positive affect before and after a lesson taught in a green environment versus a lesson taught in a classroom (p=0.234). Students with higher emotional difficulties were found to have a statistically significant larger increase in positive affect after the school lesson in greenness than after the classroom lesson (p=0.01), and a larger decrease in negative affect in greenness, although this difference did not reach statistical significance (p=0.09). The positive and negative affect of students with lower emotional difficulties did not vary as a function of the lesson environment. The third study (90) did not find a significant difference in perceived enjoyment between students taking part in a PE lesson in greenness versus the standard school grounds (p=0.66).

##### b) Social wellbeing outcomes

###### i.) Prosocial behaviour (n=4 studies)

The summary effect direction for the four studies (88, 91-93) reviewing the association between learning in nature and prosocial behaviour was of conflicting findings, with only one study reporting significant findings. A controlled before-and-after study in Denmark (92)assessed the effects of teaching 20% (one day per week) of school lessons in a forest over a three-year period, and found a significant difference in social relations outcomes (p < 0.001) between forest and school settings, however this was a very small study with only 13 respondents.

##### c) Psychological wellbeing outcomes

###### i) Attention/attention restoration (n=3 studies)

The summary effect direction for the three studies in this category was for a significant effect associated with the intervention, with two studies reporting positive outcomes that were statistically significant. A medium sized study in Italy found that primary school children performed better in tests for selective and sustained attention after a single lesson taught in the school garden, compared with a lesson taught indoors (94), and a small before-and-after study in the USA found an improved score (p < 0.001)for the ability to attend and engage after four weeks of a daily 4-hour programme of preschool education in a natural environment (93).

###### ii) Executive function skills

The two studies in this category reported a positive effect of the intervention, which was statistically significant, and the summary effect direction was therefore positive. The US study investigating impacts of teaching preschool children outdoors (93) found improved problem-solving scores (p < 0.001) after the intervention, and a Canadian study (95) found an association (p < 0.001) between performance in a self-regulation task and taking part in daily outdoor preschool lessons for a minimum of 30 minutes daily, compared to a maximum of 60 minutes weekly.

###### iii.) Resilience (n=1 study)

A single study in this category (93) found a statistically significant association between a preschool outdoor education programme and improved scores on resilience dimensions (ps < 0.05).

#### 5) Forest school

##### a) Emotional wellbeing outcomes

###### i) Emotional wellbeing scale outcomes (n=1 study)

A moderately sized (n=123 students) before-and-after study with a crossover design (96) found no significant difference in change in WEMBWS outcomes after participation in forest school versus usual school lessons (p=0.78). However, the study reported a significant difference for males only (p=0.025). The authors hypothesised that this might have been as a result of boys interacting differently with the natural environment, or having a lower life satisfaction at the baseline.

###### ii) Positive affect (n=1 study)

A small (n=18 students) before-and-after study with a crossover design (97) reported a significant effect on some dimensions of mood of participation in forest school for one day, versus normal school lessons. Energy increased significantly after forest school versus standard school (p=0.007). There was a borderline beneficial effect on stress (p=0.52), and a significant beneficial effect on hedonic tone (p=0.007) and anger (p=0.02), with the poor behaviour group (n=12 students) benefiting more than the good behaviour group.

##### b) Social wellbeing outcomes

###### i) Prosocial behaviour (n=1 study)

A controlled before-and-after study (98) found that daily attendance at a full-day nature kindergarten was associated with a significant effect on outcomes on a social skills rating scale, compared to attendance at a standard kindergarten (ps<0.01 for teacher ratings).

##### c) Psychological wellbeing outcomes

###### i) Attention/attention restoration (n=1 study)

A single study in this category (98) found no difference in outcomes for directed attention in a small Canadian study of the effects of attendance at nature kindergarten.

###### ii) Executive function skills (n= 2 studies)

Neither of the two studies in this category (98, 99) found a difference in executive function skills outcomes between children attending a nature-based preschool versus those attending a standard preschool.

###### iii) Resilience (n=2 studies)

The two studies in this category (96, 99) reported statistically significant positive effects associated with the intervention, leading to a positive and statistically significant summary effect direction. A US before-and-after study (99) found a significant increase in resilience ratings in children attending the nature preschool, but no increase in the non-nature preschool. A UK study found that secondary school student attending forest school one day per week had a significant increase in resilience scores (p<0.015), which did not occur in the control group.

## Conclusions and discussion

This review aimed to understand whether five different categories of green space intervention in educational settings impact on children’s mental wellbeing, which was defined as encompassing emotional, social and psychological wellbeing.

Based on generally very low certainty of evidence, findings from effect direction plots indicated positive associations between environmental education and improved outcomes for wellbeing scales, mood and health-related quality of life. There were insufficient studies for social and psychological wellbeing outcomes of environmental education to derive a summary effect direction.

Positive associations were also found for improved outcomes on emotional wellbeing scales after breaks or playtime in a green environment, but findings on self-esteem lacked statistical significance, and there were insufficient studies for the other emotional wellbeing categories to derive a summary effect direction. Findings from the effect direction plots indicated a positive association between pro-social behaviour and play in nature-rich playgrounds, whilst there was insufficient evidence for an association between frequency of bullying and gang activities and this intervention category. Effect direction plots also indicated a positive association between green play and attention scores, with insufficient studies investigating executive function skills and resilience.

There were insufficient or no studies investigating effects of school gardening on emotional wellbeing outcomes to be able to derive summary effect directions. Findings from effect direction plots suggested conflicting findings for the effect of school gardening on pro-social behaviour, and there were no studies at all exploring its effect on psychological wellbeing outcomes.

For studies investigating “learning in nature” interventions, effect direction plots suggested results were not statistically significant for emotional wellbeing scale and mood outcomes, with insufficient or no studies for self-esteem and quality of life outcomes. For studies exploring the effect of school gardening on social wellbeing outcomes, effect direction plots indicated conflicting findings. Positive associations were indicated by effect direction plots for attention and executive function skills outcomes after learning in nature, with insufficient studies for resilience outcomes.

Lastly, for studies exploring effects of forest school, there were insufficient or no studies on emotional and social wellbeing outcomes, and it was not possible to arrive at a summary effect direction. For psychological wellbeing outcomes associated with forest school, there were insufficient studies investigating attention outcomes, and effect direction plots suggested no clear effect for executive function skills outcomes, whilst a positive association was indicated by the effect direction plot for resilience measures and forest school.

Of the 36 studies identified which reported mental wellbeing outcomes across the three dimensions of mental wellbeing, 16 employed a before-and-after design with a separate control group, 13 studies were before-and-after studies with a cross over design and seven employed a before-and-after design without controls. Studies had a relatively uneven geographical spread over the high-income, temperate latitude countries included, with the majority located in the UK and USA. This may have been the case due to non-English language publications being excluded. Most studies had small to medium sample sizes (<300 individuals in the intervention group) and ten studies had large sample sizes. Of the 37 reports of studies, only two were rated as strong overall, ten were rated as moderate, with the remainder rated as weak. Data collection methods tended to be valid and reliable, and studies performed relatively well on study design and data collection, but they scored poorly on other assessment categories (blinding, managing confounding, reporting dropouts and selection bias).

Weaknesses identified in studies were often linked to specific features in study design. For example, a before and after study (89) in UK primary schools used a complex intervention consisting of multiple components (physical exploration of the natural environment, followed by more structured artist-led activities). Without controlling for these individual components, it is difficult to attribute changes in children’s wellbeing to the nature exposure specifically. Another weakness mentioned by authors was that the duration of the intervention may have been insufficient to produce a significant effect (67, 72, 73, 76, 96, 100), or that the intervention was diluted due to lack of funding (74, 78). Some authors questioned whether the instruments used to measure wellbeing were sensitive enough to change (68, 86, 101). A further weakness of instruments used with young children was that they might have suffered participant fatigue due to repeated daily use of very simple instruments such as a “Faces Survey” (91). Several authors raised the possibility of social desirability and observer bias (77) as observations and measurements were often taken by teachers or the study investigators, rather than independent observers, or students rated themselves (75). The majority of studies introduced a control situation by having a cross-over design (with the risk of a carry-over effect), and two studies had parallel control groups. These were not necessarily well matched, for example the experimental and control schools in an Austrian study (74) were different school types. In other studies, the equivalence of lessons given in different environments was not assessed or known to be poor (85, 90, 94, 97). Authors also highlighted that there may have been contamination bias due to exposure to natural settings outside school, and that findings from studies might not be generalisable to other settings, in particular from urban to rural settings (78).

In summary, based on summary effect directions, there was some evidence for the positive effects of environmental education and breaks in green spaces on emotional wellbeing outcomes; for beneficial effects of breaks in green spaces on social wellbeing outcomes; and for improved outcomes in particular categories of psychological wellbeing associated with breaks in green spaces, learning in nature and forest school.

### Strengths and limitations of the review

This is the first comprehensive systematic review aiming to assess the effects of green space interventions in educational settings, across all ages from 0-18, on outcomes across the three dimensions of mental wellbeing. The protocol was registered to PROSPERO and a thorough search of seven databases, as well as websites and theses conducted, and updated in March 2024. The search strategy was peer reviewed under PRESS guidance, and full text articles were assessed independently by a second reviewer where there was doubt as to their inclusion. In spite of these strengths, there were some limitations that were partially mitigated.

Title and abstract screening could not be completed in duplicate, and not all full papers received duplicate screening, however the second author (SC) reviewed full papers where there was any uncertainty about meeting inclusion criteria and a discussion-based consensus reached. We excluded studies in which interventions were targeted at children with mental health diagnoses, special educational needs or other diagnoses, and there is therefore a gap in the findings, and potential for a further review, to determine whether children in these groups would benefit to a greater extent from nature interventions.

It is possible that the EPHPP tool used to assess quantitative studies, which was designed for clinical intervention studies, tended to provide weak ratings in a field where it is difficult to adhere to a gold standard of research practice.

### Recommendations for future research

To enable us to improve the evidence base, research needs to improve study design. Most of the studies reviewed were rated moderate or weak on study design, with small or medium sample sizes (<300), and control groups often insufficiently powered. More than half of the studies included did not have separate control groups, utilising either a cross-over design or no control group at all. The review also found that many studies used a small exposure dose i.e. interventions either did not have “enough nature” or were very short-lived. Future studies therefore need to ensure there is sufficient exposure dose and duration.

A further area for improvement is around the nuance of the nature intervention. Exposures need to be more nuanced with regard to biodiversity, seasons, greenness and the nature of children’s interactions with the natural environment., and authors need to ensure that they carefully describe duration, frequency, “concentration” and character of the nature exposure. Longitudinal follow up of children post-intervention should also be strengthened, with most studies discussed here either having no follow up, or only short-term follow up over a few weeks.

## Supporting information

Supplemental File A Prisma Checklist

Supplemental file B Sample Search Strategy

Supplemental file C Summary Table

Supplemental file D Results Table

Supplemental file E Quality Appraisal

## Data Availability

All data produced in the present study are available upon reasonable request to the authors.

## References

1. Innocenti U. Worlds of Influence: Understanding what shapes child well-being in rich countries. Innocenti Report Card 16. Florence: UNICEF; 2020.

2. Rights OotHCfH. Convention on the Rights of the Child. Geneva: United Nations; 1989 20.11.1989. Contract No.: General Assembly resolution 44/25.

3. Clarke T. Children’s wellbeing and their academic achievement: The dangerous discourse of ‘trade-offs’ in education. Theory and research in education. 2020;18(3):263–94.

4. Suhrcke M, de Paz Nieves C. The impact of health and health behaviours on educational outcomes in high-income countries: a review of the evidence. International Conference on Education, Social Capital and Health; Oslo2010.

5. National Research C, Institute of Medicine Committee on Integrating the Science of Early Childhood D. In: Shonkoff JP, Phillips DA, editors. From Neurons to Neighborhoods: The Science of Early Childhood Development. Washington (DC): National Academies Press (US)

6. Copyright 2000 by the National Academy of Sciences. All rights reserved.; 2000.

6. Cowie H, Myers CA. The impact of the COVID-19 pandemic on the mental health and well-being of children and young people. Child Soc. 2021;35(1):62–74.

7. Andrew A, Cattan S, Costa Dias M, Farquharson C, Kraftman L, Krutikova S, et al. Inequalities in Children’s Experiences of Home Learning during the COVID-19 Lockdown in England. Fiscal studies. 2020;41(3):653–83.

8. Jones B, Woolfenden S, Pengilly S, Breen C, Cohn R, Biviano L, et al. COVID -19 pandemic: The impact on vulnerable children and young people in Australia. Journal of Paediatrics and Child Health. 2020;56(12):1851–5.

9. Lovell R, White MP, Wheeler B, Taylor T, Elliott L. A rapid scoping review of health and wellbeing evidence for the Framework of Green Infrastructure Standards. London/Exeter: European Centre for Environment and Human Health University of Exeter Medical School. For: Natural England/Public Health England/Ministry for Housing, Communities and Local Government, England; 2020.

10. Douglas O, Lennon M, Scott M. Green space benefits for health and well-being: A life-course approach for urban planning, design and management. Cities. 2017;66:53–62.

11. Weeland J, Moens MA, Beute F, Assink M, Staaks JPC, Overbeek G. A dose of nature: Two three-level meta-analyses of the beneficial effects of exposure to nature on children’s self-regulation. Journal of Environmental Psychology. 2019;65.

12. Mygind L, Kjeldsted E, Hartmeyer R, Mygind E, Bølling M, Bentsen P. Mental, physical and social health benefits of immersive nature-experience for children and adolescents: A systematic review and quality assessment of the evidence. Health & Place. 2019;58:102136.

13. Chawla L. Benefits of Nature Contact for Children. Journal of Planning Literature. 2015;30(4):433–52.

14. Jimenez M, DeVille N, Elliott E, Schiff J, Wilt G, Hart J, et al. Associations between Nature Exposure and Health: A Review of the Evidence. International Journal of Environmental Research and Public Health. 2021;18.

15. Vanaken G-J, Danckaerts M. Impact of Green Space Exposure on Children’s and Adolescents’ Mental Health: A Systematic Review. International Journal of Environmental Research and Public Health. 2018;15(12):2668.

16. Tillmann S, Tobin D, Avison W, Gilliland J. Mental health benefits of interactions with nature in children and teenagers: a systematic review. J Epidemiol Community Health. 2018;72(10):958–66.

17. Turner WR, Nakamura T, Dinetti M. Global Urbanization and the Separation of Humans from Nature. BioScience. 2004;54(6):585–90.

18. IPBES. Summary for policymakers of the global assessment report on biodiversity and ecosystem services of the Intergovernmental Science-Policy Platform on Biodiversity and Ecosystem Services.. Bonn, Germany: IPBES secretariat; 2019.

19. Hand KL, Freeman C, Seddon PJ, Recio MR, Stein A, van Heezik Y. Restricted home ranges reduce children’s opportunities to connect to nature: Demographic, environmental and parental influences. Landscape and Urban Planning. 2018;172:69–77.

20. de Vries S, Buijs AE, Snep RPH. Environmental Justice in The Netherlands: Presence and Quality of Greenspace Differ by Socioeconomic Status of Neighbourhoods. Sustainability. 2020;12(15):5889.

21. Schüle SA, Gabriel KMA, Bolte G. Relationship between neighbourhood socioeconomic position and neighbourhood public green space availability: An environmental inequality analysis in a large German city applying generalized linear models. International Journal of Hygiene and Environmental Health. 2017;220(4):711–8.

22. Hoffimann E, Barros H, Ribeiro AI. Socioeconomic Inequalities in Green Space Quality and Accessibility—Evidence from a Southern European City. International Journal of Environmental Research and Public Health. 2017;14(8):916.

23. Bowler DE, Buyung-Ali LM, Knight TM, Pullin AS. A systematic review of evidence for the added benefits to health of exposure to natural environments. BMC Public Health. 2010;10(1):456.

24. Twohig-Bennett C, Jones A. The health benefits of the great outdoors: A systematic review and meta-analysis of greenspace exposure and health outcomes. Environmental research. 2018;166:628–37.

25. Browning MHEM, Rigolon A. School Green Space and Its Impact on Academic Performance: A Systematic Literature Review. International journal of environmental research and public health. 2019;16(3).

26. McCormick R. Does Access to Green Space Impact the Mental Well-being of Children: A Systematic Review. Journal of Pediatric Nursing. 2017;37:3–7.

27. Mnich C, Weyland S, Jekauc D, Schipperijn J. Psychosocial and Physiological Health Outcomes of Green Exercise in Children and Adolescents-A Systematic Review. Int J Environ Res Public Health. 2019;16(21).

28. Norwood MF, Lakhani A, Fullagar S, Maujean A, Downes M, Byrne J, et al. A narrative and systematic review of the behavioural, cognitive and emotional effects of passive nature exposure on young people: Evidence for prescribing change. Landscape and Urban Planning. 2019;189:71–9.

29. Ohly H, Gentry S, Wigglesworth R, Bethel A, Lovell R, Garside R. A systematic review of the health and well-being impacts of school gardening: synthesis of quantitative and qualitative evidence. BMC Public Health. 2016;16:286.

30. Adams S, Savahl S. Nature as children’s space: A systematic review. The Journal of Environmental Education. 2017;48(5):291–321.

31. Bikomeye JC, Balza J, Beyer KM. The Impact of Schoolyard Greening on Children’s Physical Activity and Socioemotional Health: A Systematic Review of Experimental Studies. International journal of environmental research and public health. 2021;18(2):535.

32. Dankiw KA, Tsiros MD, Baldock KL, Kumar S. The impacts of unstructured nature play on health in early childhood development: A systematic review. PLOS ONE. 2020;15(2):e0229006.

33. van den Bogerd N, Dijkstra SC, Koole SL, Seidell JC, de Vries R, Maas J. Nature in the indoor and outdoor study environment and secondary and tertiary education students’ well-being, academic outcomes, and possible mediating pathways: A systematic review with recommendations for science and practice. Health & Place. 2020;66.

34. Putra IGNE, Thomas A-B, Dylan PC, Stewart AV, Eme Eseme J, Xiaoqi F. The Relationship Between Green Space and Prosocial Behaviour Among Children and Adolescents: A Systematic Review. Frontiers in psychology. 2020;11.

35. Dankiw KA, Kumar S, Baldock KL, Tsiros MD. Parent and early childhood educator perspectives of unstructured nature play for young children: A qualitative descriptive study. PLoS ONE. 2023;18(6 June):e0286468.

36. Roberts A, Hinds J, Camic PM. Nature activities and wellbeing in children and young people: a systematic literature review. Journal of Adventure Education and Outdoor Learning. 2019:1–21.

37. The effect of exposure to nature on children’s psychological well-being: a systematic review of the literature. Urban Forestry & Urban Greening. 2023;81(85).

38. Johnstone A, Martin A, Cordovil R, Fjortoft I, Iivonen S, Jidovtseff B, et al. Nature-Based Early Childhood Education and Children’s Social, Emotional and Cognitive Development: A Mixed-Methods Systematic Review. International Journal of Environmental Research & Public Health [Electronic Resource]. 2022;19(10):13.

39. Ulrich RS. Natural versus urban scenes: Some psychophysiological effects. Environment and Behavior. 1981;13:523–56.

40. Wilson E. Biophilia. Cambridge, MA: Harvard University Press; 1984.

41. Kaplan S. The Restorative Benefits of Nature: Toward an integrative framework. Journal of Environmental Psychology. 1995;15:169–82.

42. Kaplan R, Kaplan S. The experience of nature: A psychological perspective. Cambridge, MA: Cambridge University Press; 1989.

43. Kuo M. How might contact with nature promote human health? Promising mechanisms and a possible central pathway. Front Psychol. 2015;6.

44. Komori T, Fujiwara R, Tanida M, Nomura J, Yokoyama M. Effects of Citrus Fragrance on Immune Function and Depressive States. Neuroimmunomodulation 1995;2:174–80.

45. Hagerhall C, Laike T, Kuller M, Marcheschi E, Boydston C, Taylor R. Human physiological benefits of viewing nature: EEG responses to exact and statistical fractal patterns. Nonlinear dynamics, psychology and life sciences. 2015;19:1–12.

46. aan het Rot M, Moskowitz D, Young S. Exposure to bright light is associated with positive social interaction and good mood over short time periods: a naturalistic study in mildly seasonal people. Journal of Psychiatric Research. 2008;42(3):311–9.

47. Alvarsson J, Wiens S, Nilsson M. Stress recovery during exposure to nature sound and environmental noise. International Journal of Environmental Research and Public Health. 2010;7:1036–46.

48. O’Brien ME, Anderson H, Kaukel E, O’Byrne K, Pawlicki M, Von Pawel J, et al. SRL172 (killed Mycobacterium vaccae) in addition to standard chemotherapy improves quality of life without affecting survival, in patients with advanced non-small-cell lung cancer: phase III results. Annals of Oncology. 2004;15(6):906–14.

49. Richardson M, Hunt A, Hinds J, Bragg R, Fido D, Petronzi D, et al. A Measure of Nature Connectedness for Children and Adults: Validation, Performance and Insights. Sustainability. 2019;11.

50. Martin L, White MP, Hunt A, Richardson M, Pahl S, Burt J. Nature contact, nature connectedness and associations with health, wellbeing and pro-environmental behaviours. Journal of Environmental Psychology. 2020;68.

51. Arola T, Aulake M, Ott A, Lindholm M, Kouvonen P, Virtanen P, et al. The impacts of nature connectedness on children’s well-being: Systematic literature review. Journal of Environmental Psychology. 2023;85:101913.

52. Kraut R. Aristotle on well-being. The Routledge Handbook Of Philosophy Of Well-Being: Routledge; 2015.

53. Kahnemann D, Diener E, Schwarz Ne. Well-Being: the foundations of hedonic psychology. New York, NY: Russell Sage Foundation; 1999.

54. Waterman A. Two Conceptions of Happiness: Contrasts of Personal Expressiveness (Eudaimonia) and Hedonic Enjoyment. Journal of Personality and Social Psychology. 1993;64.

55. Henderson L, Knight T. Integrating the hedonic and eudaimonic perspectives to more comprehensively understand wellbeing and pathways to wellbeing. International Journal of Wellbeing. 2012;2(3):196–221.

56. Sen A. Capability and Well-being. In: Nussbaum M, Sen A, editors. The Quality of Life. Oxford, UK: Clarendon Press; 1993. p. 30–53.

57. Seligman ME, Csikszentmihalyi M. Positive psychology. An introduction. Am Psychol. 2000;55(1):5–14.

58. de Cates A, Stranges S, Blake A, Weich S. Mental well-being: An important outcome for mental health services? British Journal of Psychiatry. 2015;207(3):195–7.

59. Ferguson M, Roberts HE, McEachan RRC, Dallimer M. Contrasting distributions of urban green infrastructure across social and ethno-racial groups. Landscape and urban planning. 2018;175:136–48.

60. Mansfield KL, Ukoumunne OC, Blakemore SJ, Montero-Marin J, Byford S, Ford T, et al. Missing the context: The challenge of social inequalities to school-based mental health interventions. JCPP advances. 2023;3(2):e12165-n/a.

61. McGowan J, Sampson M, Salzwedel DM, Cogo E, Foerster V, Lefebvre C. PRESS Peer Review of Electronic Search Strategies: 2015 Guideline Statement. Journal of Clinical Epidemiology. 2016;75:40–6.

62. Thomas BH, Ciliska D, Dobbins M, Micucci S. A process for systematically reviewing the literature: providing the research evidence for public health nursing interventions. Worldviews Evid Based Nurs. 2004;1(3):176–84.

63. Campbell M, McKenzie JE, Sowden A, Katikireddi SV, Brennan SE, Ellis S, et al. Synthesis without meta-analysis (SWiM) in systematic reviews: reporting guideline. BMJ. 2020;368:n6890.

64. Thomson HJ, Thomas S. The effect direction plot: visual display of non-standardised effects across multiple outcome domains. Res Synth Methods. 2013;4(1):95–101.

65. Armijo-Olivo S, Stiles CR, Hagen NA, Biondo PD, Cummings GG. Assessment of study quality for systematic reviews: a comparison of the Cochrane Collaboration Risk of Bias Tool and the Effective Public Health Practice Project Quality Assessment Tool: methodological research. J Eval Clin Pract. 2012;18(1):12–8.

66. Harvey DJ, Montgomery LN, Harvey H, Hall F, Gange AC, Watling D. Psychological benefits of a biodiversity-focussed outdoor learning program for primary school children. Journal of Environmental Psychology Vol 67, 2020, ArtID 101381. 2020;67.

67. Pirchio S, Passiatore Y, Panno A, Cipparone M, Carrus G. The Effects of Contact With Nature During Outdoor Environmental Education on Students’ Wellbeing Connectedness to Nature and Pro-sociality. Frontiers in Psychology. 2021;12:648458.

68. Sheldrake R, Amos R, Reiss M. Children and Nature. A Research Evaluation for the Wildlife Trusts. London: The Wildlife Trusts/University College London; 2019.

69. Montgomery LN, Gange AC, Watling D, Harvey DJ. Children’s perception of biodiversity in their school grounds and its influence on their wellbeing and resilience. Journal of Adventure Education and Outdoor Learning. 2022.

70. Barrable A, Booth D, Adams D, Beauchamp G. Enhancing Nature Connection and Positive Affect in Children through Mindful Engagement with Natural Environments. International Journal of Environmental Research & Public Health [Electronic Resource]. 2021;18(9):30.

71. Dopko RL, Capaldi CA, Zelenski JM. The psychological and social benefits of a nature experience for children: A preliminary investigation. Journal of Environmental Psychology. 2019;63:134–8.

72. Sprague N, Berrigan D, Ekenga CC. An Analysis of the Educational and Health-Related Benefits of Nature-Based Environmental Education in Low-Income Black and Hispanic Children. Health equity. 2020;4(1):198–210.

73. Sprague NL, Ekenga CC. The impact of nature-based education on health-related quality of life among low-income youth: results from an intervention study. Journal of Public Health. 2021;44(2):394–401.

74. Kelz C, Evans GW, Röderer K. The Restorative Effects of Redesigning the Schoolyard. Environment and Behavior. 2015;47(2):119–39.

75. Wallner P, Kundi M, Arnberger A, Eder R, Allex B, Weitensfelder L, et al. Reloading Pupils’ Batteries: Impact of Green Spaces on Cognition and Wellbeing. Int J Environ Res Public Health. 2018;15(6).

76. Barton J, Sandercock G, Pretty J, Wood C. The effect of playground- and nature-based playtime interventions on physical activity and self-esteem in UK school children. International journal of environmental health research. 2015;25(2):196–206.

77. Carrus G, Passiatore Y, Pirchio S, Scopelliti M. Contact with nature in educational settings might help cognitive functioning and promote positive social behaviour/El contacto con la naturaleza en los contextos educativos podría mejorar el funcionamiento cognitivo y fomentar el comportamiento social positivo. Psyecology: Revista Biling/Bilingual Journal of Environmental Psychology. 2015;252.

78. van Dijk-Wesselius JE, Maas J, Hovinga D, van Vugt M, van den Berg AE. The impact of greening schoolyards on the appreciation, and physical, cognitive and social-emotional well-being of schoolchildren: A prospective intervention study. Landscape and Urban Planning. 2018;180:15–26.

79. Bates CR, Bohnert AM, Gerstein DE. Green Schoolyards in Low-Income Urban Neighborhoods: Natural Spaces for Positive Youth Development Outcomes. Frontiers in Psychology. 2018;9.

80. Brussoni M, Ishikawa T, Brunelle S, Herrington S. Landscapes for play: Effects of an intervention to promote nature-based risky play in early childhood centres. Journal of Environmental Psychology. 2017;54:139–50.

81. Raney MA, Hendry CF, Yee SA. Physical Activity and Social Behaviors of Urban Children in Green Playgrounds. American Journal of Preventive Medicine. 2019;56(4):522–9.

82. Raney MA, Bowers AL, Rissberger AL. Recess Behaviors of Urban Children 16 Months After a Green Schoolyard Renovation. Journal of Physical Activity & Health. 2021;18(5):563–70.

83. Amicone G, Petruccelli I, De Dominicis S, Gherardini A, Costantino V, Perucchini P, et al. Green Breaks: The Restorative Effect of the School Environment’s Green Areas on Children’s Cognitive Performance. Frontiers in Psychology. 2018;9.

84. Anabitarte A, García-Baquero G, Andiarena A, Lertxundi N, Urbieta N, Babarro I, et al. Is brief exposure to green space in school the best option to improve attention in children? International journal of environmental research and public health. 2021;18(14):7484.

85. Pollin S, Retzlaff-Furst C. The school garden: A social and emotional place. Frontiers in Psychology. 2021;12.

86. Block K, Gibbs L, Staiger PK, Gold L, Johnson B, Macfarlane S, et al. Growing community: The impact of the Stephanie Alexander Kitchen Garden Program on the social and learning environment in primary schools. Health Education & Behavior. 2012;39(4):419–32.

87. Waliczek TM, Bradley JC, J.M./Z. The Effect of School Gardens on Children’s Interpersonal Relationships and Attitudes Toward School. HortTech. 2001;11(3).

88. van den Berg AE, Hovinga D, Joven M, Steensma R, Maas J. Strengthening the pedagogical use of the outdoor area at nature-based daycare centers: An intervention study. Urban Forestry & Urban Greening. 2024;92:N.PAG-N.PAG.

89. Moula Z, Walshe N, Lee E. “It was like I was not a person, it was like I was the nature”: The impact of arts-in-nature experiences on the wellbeing of children living in areas of high deprivation. Journal of Environmental Psychology. 2023;90:1–12.

90. Reed K, Wood C, Barton J, Pretty JN, Cohen D, Sandercock GRH. A Repeated Measures Experiment of Green Exercise to Improve Self-Esteem in UK School Children. PLOS ONE. 2013;8(7):e69176.

91. Largo-Wight E, Guardino C, Wludyka PS, Hall KW, Wight JT, Merten JW. Nature contact at school: The impact of an outdoor classroom on children’s well-being. International journal of environmental health research. 2018;28(6):653–66.

92. Mygind E. A comparison of childrens’ statements about social relations and teaching in the classroom and in the outdoor environment. Journal of Adventure Education and Outdoor Learning. 2009;9(2):151–69.

93. Scogin SC, D’Agostino SR, Dykstra J, Veine C, Schuen A. Exploring the effects of a short-term, nature-based preschool experience: a mixed-methods investigation. Journal of Outdoor and Environmental Education. 2023.

94. Mason L, Manzione L, Ronconi A, Pazzaglia F. Lessons in a Green School Environment and in the Classroom: Effects on Students’ Cognitive Functioning and Affect. International Journal of Environmental Research and Public Health. 2022;19(24).

95. Taylor AF, Butts-Wilmsmeyer C. Self-regulation gains in kindergarten related to frequency of green schoolyard use. Journal of Environmental Psychology Vol 70, 2020, ArtID 101440. 2020;70.

96. Roberts A. Forest School and Mental Wellbeing. Thesis. In: University CCC, editor. 2017.

97. Roe J, Aspinall P. The restorative outcomes of forest school and conventional school in young people with good and poor behaviour. Urban Forestry & Urban Greening. 2011;10(3):205–12.

98. Mueller U, Temple V, Smith B, Kerns K, Eycke K, Crane J, et al. Effects of Nature Kindergarten Attendance on Children’s Functioning. Children, Youth and Environments. 2017;27(2):47–69.

99. Ernst J, Burçak F. Young Children’s Contributions to Sustainability: The Influence of Nature Play on Curiosity, Executive Function Skills, Creative Thinking, and Resilience. Sustainability. 2019.

100. Pirchio S, Passiatore Y, Panno A, Cipparone M, Carrus G. The effects of contact with nature during outdoor environmental education on students’ wellbeing connectedness to nature and pro-sociality. Frontiers in Psychology. 2021;12.

101. Wood C, Gladwell V, Barton J. A repeated measures experiment of school playing environment to increase physical activity and enhance self-esteem in UK school children. PloS one. 2014;9(9):e108701.

