## Supplemental file B Sample Search Strategy for "Impacts of green space interventions in educational settings on children and young people’s mental wellbeing: a systematic review"

Search strategy OVID medline final 14.12.2020

Database: Ovid MEDLINE(R) and Epub Ahead of Print, In-Process & Other Non-Indexed Citations, Daily and Versions(R) <1946
to December 04, 2020>
Search Strategy:
--------------------------------------------------------------------------------
1 ((natur* or green* or wild*) adj5 (space* or environ* or setting* or element* or factor* or intervention* or
activ* or experience* or play* or ground* or schoolyard* or program* or education*)).mp. (132921)
2 (outdoor* adj5 (space* or experience* or environ* or intervention* or setting* or activit* or play* or program* or
education* or time or teaching or learn* or classroom*)).mp. (6979)
3 (natur* adj2 contact).mp. (496)
4 horticult*.mp. (3337)
5 garden*.mp. (13629)
6 allotment*.mp. (618)
7 friluftsliv.mp. (6)
8 udeskole.mp. (2)
9 ((eco or ecol*) adj5 School*).mp. (237)
10 landscape*.mp. (59262)
11 (tree* or forest school*).mp. (152853)
12 (biodivers* or bio-divers*).mp. (51191)
13 exp Gardens/ or exp Horticulture/ or exp Gardening/ (1267)
14 or/1-13 (401385)
15 (school* or educat*).ti,ab. (815661)
16 Education/ or Schools/ or Schools, Nursery/ (60506)
17 early learning.ti,ab. (993)
18 early year*.ti,ab. (4065)
19 (nurser* or kindergarten).ti,ab. (16655)
20 (preschool* or pre-school*).ti,ab. (33663)
21 or/15-20 (866387)
22 ((mental or psychological) adj5 (wellbeing or well-being or health)).ti,ab. (180796)
23 (stress* or emotion*).ti,ab. (1027577)
24 behavio$r*.ti,ab. (982014)
25 (self-esteem or selfesteem or self-confidence or selfconfidence or resilien* or coping).ti,ab. (110765)
26 (self-regulati* or selfregulati* or restorati* or relax*).ti,ab. (306218)
27 ((academic or cognitive) adj3 achieve*).mp. (6709)
28 (psychological adj2 outcome*).mp. (3872)
29 (natur* adj3 connect*).ti,ab. (1141)
30 Mental Health/ (40321)
31 Child Development/ (46836)
32 (WEMWBS or warwick-edinburgh mental wellbeing scale).mp. (161)
33 (happiness or happy or confiden* or hedonic).mp. (605975)
34 (restorative adj5 (effect* or potential or experience*)).mp. [mp=title, abstract, original title, name of
substance word, subject heading word, floating sub-heading word, keyword heading word, organism supplementary concept
word, protocol supplementary concept word, rare disease supplementary concept word, unique identifier, synonyms] (1834)

35 (psychological adj5 restorat*).mp. [mp=title, abstract, original title, name of substance word, subject heading
word, floating sub-heading word, keyword heading word, organism supplementary concept word, protocol supplementary
concept word, rare disease supplementary concept word, unique identifier, synonyms] (123)
36 (self-acceptance or selfacceptance).mp. (747)
37 (mastery or competen* or autonomy).mp. (284618)
38 (attentive* or attention span*).mp. (6116)
39 (problem solving or conflict resolution*).mp. (39251)
40 eudaimonic.mp. (278)
41 ((social or relationship*) adj3 (skills or peer or peers)).mp. (14570)
42 prosocial.mp. (4788)
43 (empath* or emotional* intelligen*).mp. (32637)
44 ((aptitude* or skill*) adj3 (learn* or play*)).mp. (9930)
45 or/22-44 (3231837)
46 14 and 21 and 45 (3872)
47 limit 46 to english language (3701)
48 47 not (Animals/ not (Animals/ and Humans/)) (3613)
