## Supplemental file C Summary Table for "Impacts of green space interventions in educational settings on children and young people’s mental wellbeing: a systematic review"

**Summary of included quantitative studies (N=37)**

| First author (year) | Study design | Country | Type of education al setting | Sample size (baseline) | Sample characteristics | Intervention and duration | Comparison or control | Intervention category | Outcomes measured (aspects of mental wellbeing) | Quality apprais al (EPHPP overall score) |
|---|---|---|---|---|---|---|---|---|---|---|
| **Amicone (2018) (1) Journal article** | Studies 1 and 2: Before-and-after study with cross-over design | Italy | Primary school | Study 1: 82 children<br>Study 2: 36 children | Ages 10-11<br>Study 1 mean age 10.1<br>52% male<br>Study 2 mean age 10.8<br>51% male | Study 1 Competitive team game in green environment in school grounds. 30 minutes duration<br>Study 2 Free play in green space. 30 minutes duration. | Study 1 Same activity as in intervention, carried out in built environment<br>Study 2 Free play in built playground area. 30 minutes duration. | Break/playtime in green environment | Study 1: Sustained and selective attention; working memory; impulse control<br>Perceived restorativeness.<br>Study 2: Sustained and selective attention. Perceived restorativeness. | Moderate |
| **Anabirtate (2021) (2)** | Before-and-after study with cross-over design | Spain | Primary school | 167 children | Mean age 7 (SD 0.3)<br>Low SE deprivation index | Game and snack in green space (60 minutes) | Game (identical to intervention) and snack in grey space (60 minutes) | Break/playtime in green environment | Attention | Moderate |
| **Barrable (2021) (3) Journal article** | Before-and-after | UK | Primary school | 74 children | 9-10 years<br>Mean age 9.51 years<br>45% girls<br>39% boys | Visit to nature reserve and participation in 3 mindfulness-based activities | No control or comparison group | Environmental education | Positive and negative affect | Weak |

**Summary of included quantitative studies (N=37)**

| | | | | | | | | | | |
|---|---|---|---|---|---|---|---|---|---|---|
| | | | | | 16% gender not reported | 2.5 hours duration | | | | |
| **Barton (2015) (4) Journal article** | Before-and-after with cross-over design | UK | Primary school | 52 children | Mean age 8.8 Gender not reported | Nature-based orienteering activity in school grounds. 55 minutes duration. | Free play on concrete playground area, with small loose pieces of equipment. 55 minutes duration. | Break/playtime in green environment | Self-esteem. | Weak |
| **Bates (2018) (5) Journal article** | Before-and-after study | USA | Elementary school, middle school | 3 schools 1518 pupils 33 school staff 64 caregivers | Ages 8-14 55% male 83.9% BAME groups 58%-96% low income students | Recent participation in playground greening through "Space to Grow" project (renovation occurred 3-6 months prior to study). 12 months duration for serial outcome measurements. | No comparison group. Measurements at three time points | Break/playtime in green environment | Positive social interactions. Teasing and bullying behaviour Presence of gangs. Perception of safety and injury numbers. | Weak |
| **Block (2012) (6) Journal article** | Controlled before-and-after | Australia | Primary school | 764 children 562 parents | 8-12 years 46% boys | Stephanie Alexander Kitchen Garden Program. 45-60 min in garden class & 90 min in kitchen | Control group received no intervention | School gardening | Child wellbeing Child cooperative behaviours | Weak |

**Summary of included quantitative studies (N=37)**

| | | | | | | | | | | |
|---|---|---|---|---|---|---|---|---|---|---|
| | | | | | | class/week for 12-25 mnths | | | | |
| **Brussoni (2017) (7) Journal article** | Before-and-after | Canada | Early childhood centre | 48 children | Ages 4-5<br>Mean age 4.3<br>53% boys<br>BAME groups 31% | Modification of outdoor play spaces according to "7 Cs" (character, context, connectivity, clarity, change, chance and challenge) criteria to increase risky and green space play opportunities | Convergent repeated measures design | Break/playtime in green environment | Strengths and Difficulties Questionnaire (emotional symptoms, conduct problems, hyperactivity, peer relationship problems, prosocial behaviours).Preschool Social Behaviour Scale (aggression, prosocial behaviour, depressed affect). | Moderate |
| **Carrus (2015) (8) Journal article** | Before-and-after with cross-over design | Italy | Early years childcare centres | 39 children | 18-36 months<br>Gender not described | Free play in the gardens of the childcare centres on 7 days | No control group. Within-subjects design. Children were observed before and after their free play on either "garden days" or "non garden days" | Break/playtime in green environment | Quality of children's affective state | Weak |

**Summary of included quantitative studies (N=37)**

| | | | | | | | | | | |
|---|---|---|---|---|---|---|---|---|---|---|
| **Dopko (2019) (9) Journal article** | Before-and-after with crossover design | Canada | Elementary school | 80 children | Ages 10-11<br>Mean age 10.5<br>50% male | Field visit to nature school with student-initiated, exploratory learning. Duration 4 hours. | Repeated measures design, no control group | Environmental education | Mood<br>Enjoyment (vitality, low-arousal pleasant affect, fun)<br>Pro-social behaviour | Weak |
| **Ernst (2019)(a) (10) Journal article**<br>**Ernst (2019) (b) (11) Journal article** | Controlled before-and-after | USA | Preschool | 92 pupils (78 intervention, 14 comparator group) | Intervention: Mean age 4<br>50% male<br>Comparison: Mean age 4<br>64% male | "Nature pre-school": pre-schools with "wild" natural settings and natural playscapes in their outdoor spaces. Time spent in free play outdoors 4-5 h daily for one school year. | "Non-nature pre-schools": Free play time mostly spent indoors, with 30-60 min daily outdoor playtime, in outdoor space containing play equipment. | Nature pre-school/forest school | Resilience sub-scales (initiative, self-regulation, attachment/relationship) | Weak |
| **Harvey (2020) (12) Journal article** | Controlled before-and-after | UK | Primary school | 456 children including controls included in analysis | Age 8-11<br>Mean age 9.06<br>37.9% identified male<br>50.7% identified female<br>11.4% did not indicate gender | Environmental education, mostly delivered in school grounds by Biological Sciences students.<br>1 hour per week, 7 | Controls (N=90) did not receive any intervention | Environmental education | Mood<br>Global wellbeing score | Weak |

**Summary of included quantitative studies (N=37)**

| | | | | | | | | | | |
|---|---|---|---|---|---|---|---|---|---|---|
| | | | | | | sessions per term over three terms. | | | | |
| **Kelz (2015) (13) Journal article** | Controlled before-and-after | Austria | Middle school | 133 students | Age 13-15<br>51.5% male | Schoolyard renovation-10 shrubs, 10 pot plants, enhanced seating, soccer goals, drinking fountain. Exposure lasted 7 weeks. | Two control schools in same street. No schoolyard in school A (students sat outside entrance); school B had outdoor area with seating on a lawn with some trees and shrubs. | Break/playtime in green environment | Global wellbeing score<br>Recovery from stress | Weak |
| **Largo-Wight (2019) (14) Journal article** | Before-and-after with crossover design | USA | Kindergarten | 37 children | Age 5-6<br>54% male<br>15% BAME groups | Teaching regular language lesson in a nature-based outdoor setting. 30 minute duration, with two classes rotating daily between intervention and control, | Teaching regular language class in the usual indoor classroom, 30 minute duration. | Learning in nature | Child engagement behaviour<br>Child focus and attention<br>Child happiness and wellbeing | Moderate |

**Summary of included quantitative studies (N=37)**

| | | | | | | | | | | |
|---|---|---|---|---|---|---|---|---|---|---|
| | | | | | | observed for 6 weeks. | | | | |
| **Mason (2022) (15) Journal article** | Before-and-after with crossover design | Italy | Primary school | 65 children 30 female Mean age = 8.25 (SD = 0.43) | Children in second and third grade of primary school | Intervention was a single lesson (1h-1h10min.) taught outdoor in the green school garden (no teacher interaction with nature) | Comparison was a single lesson taught indoors in the classroom | Learning in nature | Affective state Perceived restorativeness Selective and sustained attention | Weak |
| **Montgomery (2022) (16) Journal article** | Controlled before-and-after | United Kingdom | Primary school | 509 (intervention) 243 (control) | Intervention group mean age = 8.81 (SD = 0.68) Control group mean age = 8.94 (SD = 0.86) Gender not stated | Intervention took place across one academic year, seven sessions of one hour each week per term (21 sessions in total). Intervention consisted of presentation in classroom followed by hands-on learning about biodiversity in school grounds | Control group of children who did not participate in the biodiversity education activities | Environmental education | Wellbeing Resilience | Weak |
| **Moula (2023) (17)** | Before-and-after | United Kingdom | Primary school | 101 children (data included | Children aged 7-10 | Intervention was delivered across eight | No control group | Learning in nature | Wellbeing | Weak |

**Summary of included quantitative studies (N=37)**

| | | | | | | | | | | |
|---|---|---|---|---|---|---|---|---|---|---|
| **Journal article** | (within-subject) | | | from 95 children) | | consecutive weeks in the summer term; one full day per school week. The art-in-nature intervention consisted of children physically engaging with the natural environment at school and then making artwork with or about nature | | | | |
| **Mueller (2017) (18) Journal article** | Controlled before-and-after | Canada | Kindergarten | 86 children (43 nature kindergarten, 41 control) | Mean age 5.3 years<br>50% male | Daily attendance at full-day nature kindergarten. | Attendance at regular kindergarten, part of an elementary school | Nature preschool/forest school | Directed attention inhibition<br>Social skills and psychological health | Weak |
| **Mygind (2009) (19) Journal article** | Controlled before-and-after | Denmark | Primary school | 19 children | Age 9 at first measurement<br>Gender not reported | 20% of school lessons taught in a forest. Duration 1 day per week over 3 years. | Normal classroom environment | Learning in nature | Social relations | Weak |
| **Pirchio (2021) (20)** | Controlled before- | Italy | **Studies 1 and 2:** Primary school; | 407 total (intervention group 154; | 54.1% males<br>246 students aged 9-10 | 3 outdoor visits to nature reserve for students and | Control group of children not participating | Environmental education | Psycho-physical wellbeing; life satisfaction | Weak |

**Summary of included quantitative studies (N=37)**

| | | | | | | | | | | |
|---|---|---|---|---|---|---|---|---|---|---|
| **Journal article** | and-after | | secondary school (1st year) | control group 253) | 161 students aged 11 | teachers; one visit per month; 4th visit with parents. 4 different environmental education workshops for all students . Maximum 4 hours per visit. | in intervention , matched to intervention group | | | |
| | | | | 338 students (intervention group 170; control group 168) | 48.8% males 171 students aged 9-10 167 studnets aged 11 | 4 outdoor visits to nature reserve as for study 1 (different site from study 1) Environmental education activities varied from study 1. | | | | |
| **Pollin (2021) (21) Journal article** | Before-and-after with crossove r design | Germany | Secondary school | 53 students | 22 boys 31 girls 6th grade Ages 12-13 | Biology lessons (teaching curriculum module "Plants and Soil" ) Intervention over 10 weeks, changing lessons weekly between classroom and garden. | Comparison was science lessons in classroom | School gardening | "Emotions" | Weak |

**Summary of included quantitative studies (N=37)**

| | | | | | | | | | | |
|---|---|---|---|---|---|---|---|---|---|---|
| | | | | | | Outdoor lessons consisted of student-led gardening with teacher support. | | | | |
| **Raney (2019) (22) Journal article** | Controlled before-and-after | USA | Elementary (primary) school | 306 children at first data collection point | Grades 1-5 of US elementary school<br>45.8% male | Conversion of asphalt playground to a greened playground over summer holidays | Asphalt playground in control school maintained | Break/playtime in green environment | Social interactions (physical and verbal conflict rates, minutes spent alone, minutes spent in small groups). SOCARP tool. | Moderate |
| **Raney (2021) (23) Journal article** | Controlled before-and-after | USA | Elementary school | 93 children in control group<br>237 in experimental group (note this is the continuation of the study reported in (22)) | Grades 1-6 of US elementary school (ages 5-11)<br>2 schools in Los Angeles<br>Majority socioeconomically disadvantaged<br>Majority Hispanic<br>53 girls and 40 boys in control group<br>115 girls and 122 boys in experimental group | 21,000 square feet of asphalt at the experimental location was replaced by green space (trees, mulch, boulders, grass, outdoor classroom.<br>Observations made prior to, just after, 4 months after, and 16 months after greening measure | Asphalt playground in control school maintained | Break/playtime in green environment | Social interactions | Moderate |

**Summary of included quantitative studies (N=37)**

| | | | | | | | | | | |
|---|---|---|---|---|---|---|---|---|---|---|
| **Reed (2013) (24) Journal article** | Before-and-after with crossover design | UK | Secondary school | 86 children | Mean age 11.4<br>Gender not reported | 1.5 mile run of a single lap course through the local country park. Duration: 1 physical education lesson | 1.5 mile run of a one lap course around the school campus. | Learning in nature | Self esteem (Rosenberg self esteem scale) and enjoyment | Moderate |
| **Roberts 2017 (25) Thesis** | Before-and-after with crossover design | UK | Secondary school | 130 students | Age 11-12<br>53.9% male | Participation in forest school. Duration 1 day per week over 5 weeks | Non-participation in forest school (crossover design) | Nature preschool/forest school | Wellbeing (global)- WEMWBS<br>Resilience (Sense of Mastery scale) | Moderate |
| **Roe (2011) (26) Journal article** | Before-and-after with crossover design | UK | Secondary school (one mainstream, one residential) | 18 students | Age 11-12<br>83.3% male | Participation in forest school near the student's usual school. Duration 5 hours on one school day. | Attendance at usual school (crossover design) | Nature preschool/forest school | Mood Adjective Checklist (MACL) - hedonic tone, energy, stress, anger<br>Project planning and affective dimensions on personal project scale | Weak |
| **Scogin (2023)(27) Journal article** | Before-and-after | USA | Pre-school | 69 children (data available for 45-62 children depending on element of rating scale tested) | Age 4-5<br>58% male<br>50% White<br>9.7% Hispanic<br>1.6% African American<br>6.5% Asian<br>32.3% Multiracial | Four hours per day for 4 weeks<br>Intervention consisted of 4-hour programme (indoor and outdoor play, hiking, small group work and whole group | No control | Learning in nature | Social-emotional outcomes | Weak |

**Summary of included quantitative studies (N=37)**

| | | | | | | | | | | |
|---|---|---|---|---|---|---|---|---|---|---|
| | | | | | | meetings. Activities and learning were related to an insect-based curriculum. | | | | |
| **Sheldrake (2019) (28) Report** | Before-and-after | UK | Primary school | 451 children | Age 7-11<br>48% boys | Environmental education activities at Wildlife Trust sites. Duration range from short ( half day) to medium (3 days over 3 weeks) to long (6 days over 6 weeks) | No control group (pre test-post test design) | Environmental education | Subjective wellbeing | Weak |
| **Sprague (2020) (29) Journal article** | Before-and-after | USA | Elementary and middle school | 122 students | Mean age 11.9<br>50% boys<br>86.7% from BAME groups | Monthly nature-based field trips accompanying programme of classroom-based environmental education. Duration 1 day per month outdoors. | No control group (pre test-post test design) | Environmental education | HRQoL domains | Weak |
| **Sprague (2021) (30) Journal article** | Controlled before- | USA | Elementary and middle school | 362 students (297 intervention, 65 control) | 86% of schools had student body that was 95% or more | 15-week curriculum based around environment | Control group did not receive intervention | Environmental education | HRQoL domains | Weak |

**Summary of included quantitative studies (N=37)**

| | | | | | | | | | | |
|---|---|---|---|---|---|---|---|---|---|---|
| | and-after | | | | eligible for free school lunches<br>Mean age overall 11.9 (intervention group 11.4, control group 12.7)<br>Gender 52.2% male<br>47.8% female<br>90.7% Black ethnicity, 9.3% other<br>No significant differences between control and intervention only for age | and health topics. Partly classroom based; included 5 field trips between 3h-26h duration. | and followed standard curriculum | | | |
| **Taylor (2020) (31) Journal article** | Controlled before-and-after | Canada | Kindergarten | Time period 1 (T1) 135 children<br>Time period 2, T2, 250 children | Mean age T1 = 5.1<br>T2= 4.3<br>T1 53.3% boys<br>T2 54.4% boys | Minimum 30 minutes daily (60 minutes daily at T2) of curriculum taught in outdoor greenspace daily. Duration 12 weeks (T1) and 9 weeks (T2) | Maximum of 60 minutes per week of curriculum taught in outdoor greenspace. | Learning in nature | Behavioural self-regulation (Child Behaviour Rating Scale, Head-Toes-Knees-Shoulders Task) | Strong |

**Summary of included quantitative studies (N=37)**

| | | | | | | | | | | |
|---|---|---|---|---|---|---|---|---|---|---|
| **Van den Berg (2024) (32) Journal article** | Controlled study with post-design | Netherlands | Daycare centres | 133 children in 6 intervention locations and 7 control locations Complete data for 96 children (play behaviour) and 111 children (wellbeing and involvement) | 0-4 years | Intervention took place over 1 year. Intervention consisted of strengthening caregiver interactive skills in relation to use of the outdoor play area and supporting them in developing programmes of outdoor activities | Matched daycare centres (matched for size, naturalness of outdoor area and urbanity) | Learning in nature | Wellbeing<br>Social behaviour<br>Involvement | Moderate |
| **Van Dijk-Wesselius (2018) (33) Journal article** | Controlled before-and-after | Netherlands | Primary school | 2,031 children | Mean age 8.6<br>48.6% boys (intervention group) | Schoolyard greening (grassy hills, shrubs, trees, garden) after baseline measurement. Duration of exposure 48 months | Paved schoolyards with some play equipment | Break/playtime in green environment | Perceived restorative quality<br>Sky Search Task<br>Social Orientation choice Card (SOCC)<br>Social behaviour scale<br>Subscale emotional functioning of paediatric QoL scale | Weak |
| **Waliczek (2001) (34) Journal article** | Controlled before-and-after | USA | Elementary and middle school | 598 students | Age 7-14<br>Gender proportion not reported | Project GREEN School gardening projects. Duration 1 term | No school gardening intervention. | School gardening | Interpersonal relationships (Self-Report of Personality Scale from BASC system) | Weak |

**Summary of included quantitative studies (N=37)**

| | | | | | | | | | | |
|---|---|---|---|---|---|---|---|---|---|---|
| **Wallner (2018) (35) Journal article** | Before-and-after with crossover design | Austria | Secondary school | 64 students | Mean age 16.6<br>50% male | Three distinct green spaces used for lunch breaks: condition 1 urban park, condition 2 larger park, condition 3 forest.<br>Duration: Lunch break, consisting of 20 min walk to site, then lunch, then brief walk, then relaxation | No control group. Crossover design | Break/playtime in green environment | Wellbeing (Nitsch self-condition scale (readiness for action, readiness for exertion, alertness, state of mood, tension/relaxation, recuperation))<br>Attention | Weak |
| **Whitburn (2023) (36) Journal article** | Controlled before-and-after | New Zealand | Primary and middle schools | 210 students participate in intervention group<br>47 students served as control group (matched for age, ethnicity,nature connectedness and pro environmental behaviour) | Age 7-13<br>38.1% female<br>Mean age 10.7<br>61.9% New Zealand European<br>24.9% Maori<br>25.3% Pacific Peoples<br>11.7% Asian<br>5.4% Other<br>(sum is more than 100% as children allowed to | Environmental education field trips to one of three destinations: a wildlife sanctuary, a zoo and a freshwater/marine environment location<br>The intervention duration was a half-day | Non-participation in environmental field trips | Environmental education | Life satisfaction<br>Vitality | Strong |

**Summary of included quantitative studies (N=37)**

| | | | | | | | | | | |
|---|---|---|---|---|---|---|---|---|---|---|
| | | | | | select more than one) | | | | | |
| **Wood (2014) (37) Journal article** | Before-and-after with crossover design | UK | Primary school | 25 students | Mean age 8.6 48% boys | Morning break and lunch break spent in green playing area (field, shrubs, trees). Duration 45 min daily for 1 week each for intervention and control | Break times spent in concrete areas of playground | Break/playtime in green environment | Self-esteem (Rosenberg Self-esteem Scale) | Moderate |

[a] Also included for qualitative studies summary


1. Amicone G, Petruccelli I, De Dominicis S, Gherardini A, Costantino V, Perucchini P, et al. Green breaks: The restorative effect of the school environment's green areas on children's cognitive performance. Frontiers in Psychology. 2018;9.
2. Anabitarte A, García-Baquero G, Andiarena A, Lertxundi N, Urbieta N, Babarro I, et al. Is brief exposure to green space in school the best option to improve attention in children? International journal of environmental research and public health. 2021;18(14):7484.
3. Barrable A, Booth D, Adams D, Beauchamp G. Enhancing nature connection and positive affect in children through mindful engagement with natural environments. International Journal of Environmental Research and Public Health. 2021;18(9):4785.
4. Barton J, Sandercock G, Pretty J, Wood C. The effect of playground- and nature-based playtime interventions on physical activity and self-esteem in UK school children. International journal of environmental health research. 2015;25(2):196-206.
5. Bates CR, Bohnert AM, Gerstein DE. Green schoolyards in low-income urban neighborhoods: Natural spaces for positive youth development outcomes. Frontiers in Psychology. 2018;9.
6. Block K, Gibbs L, Staiger PK, Gold L, Johnson B, Macfarlane S, et al. Growing community: The impact of the Stephanie Alexander Kitchen Garden Program on the social and learning environment in primary schools. Health Education & Behavior. 2012;39(4):419-32.

**Summary of included quantitative studies (N=37)**


7. Brussoni M, Ishikawa T, Brunelle S, Herrington S. Landscapes for play: Effects of an intervention to promote nature-based risky play in early childhood centres. Journal of Environmental Psychology. 2017;54:139-50.

8. Carrus G, Passiatore Y, Pircho S, Scopelliti M. Contact with nature in educational settings might help cognitive functioning and promote positive social behaviour/El contacto con la naturaleza en los contextos educativos podria mejorar el funcionamiento cognitivo y fomental el comportamiento social positivo. Psyecology: Revista Biling/Bilingual Journal of Environmental Psychology. 2015;6(2).

9. Dopko RL, Capaldi CA, Zelenski JM. The psychological and social benefits of a nature experience for children: A preliminary investigation. Journal of Environmental Psychology. 2019;63:134-8.

10. Ernst J, Burcak F. Young Children's Contributions to Sustainability: The Influence of Nature Play on Curiosity, Executive Function Skills, Creative Thinking, and Resilience. Sustainability. 2019;11(15).

11. Ernst J, Johnson M, Burcak F. The Nature and Nurture of Resilience: Exploring the Impact of Nature Preschools on Young Children's Protective Factors. International Journal of Early Childhood Environmental Education. 2019;6(2):7-18.

12. Harvey DJ, Montgomery LN, Harvey H, Hall F, Gange AC, Watling D. Psychological benefits of a biodiversity-focussed outdoor learning program for primary school children. Journal of Environmental Psychology. 2020;67.

13. Kelz C, Evans GW, Röderer K. The Restorative Effects of Redesigning the Schoolyard. Environment and Behavior. 2013;47(2):119-39.

14. Largo-Wight E, Guardino C, Wludyka PS, Hall KW, Wight JT, Merten JW. Nature contact at school: The impact of an outdoor classroom on children's well-being. International journal of environmental health research. 2018;28(6):653-66.

15. Mason L, Manzione L, Ronconi A, Pazzaglia F. Lessons in a Green School Environment and in the Classroom: Effects on Students' Cognitive Functioning and Affect. International Journal of Environmental Research and Public Health. 2022;19(24).

16. Montgomery LN, Gange AC, Watling D, Harvey DJ. Children's perception of biodiversity in their school grounds and its influence on their wellbeing and resilience. Journal of Adventure Education and Outdoor Learning. 2022.

17. Moula Z, Walshe N, Lee E. "It was like I was not a person, it was like I was the nature": The impact of arts-in-nature experiences on the wellbeing of children living in areas of high deprivation. Journal of Environmental Psychology. 2023;90:1-12.

18. Mueller U, Temple V, Smith B, Kerns K, Ten Eycke K, Crane J, et al. Effects of Nature Kindergarten Attendance on Children's Functioning. Children, Youth and Environments. 2017;27(2):47-69.

19. Mygind E. A comparison of childrens' statements about social relations and teaching in the classroom and in the outdoor environment. Journal of Adventure Education & Outdoor Learning. 2009;9(2):151-69.

20. Pirchio S, Passiatore Y, Panno A, Cipparone M, Carrus G. The effects of contact with nature during outdoor environmental education on students' wellbeing, connectedness to nature and pro-sociality. Frontiers in Psychology. 2021;12.

21. Pollin S, Retzlaff-Furst C. The school garden: A social and emotional place. Frontiers in Psychology. 2021;12.

22. Raney MA, Hendry CF, Yee SA. Physical activity and social behaviors of urban children in green playgrounds. American Journal of Preventive Medicine. 2019;56(4):522-9.

**Summary of included quantitative studies (N=37)**


23. Raney MA, Bowers AL, Rissberger AL. Recess Behaviors of Urban Children 16 Months After a Green Schoolyard Renovation. Journal of Physical Activity & Health. 2021;18(5):563-70.
24. Reed K, Wood C, Barton J, Pretty JN, Cohen D, Sandercock GRH. A Repeated Measures Experiment of Green Exercise to Improve Self-Esteem in UK School Children. PLOS ONE. 2013;8(7):e69176.
25. Roberts A. Forest School and Mental Wellbeing. Thesis. In: University CCC, editor. 2017.
26. Roe J, Aspinall P. The restorative outcomes of forest school and conventional school in young people with good and poor behaviour. Urban Forestry & Urban Greening. 2011;10(3):205-12.
27. Scogin SC, D'Agostino SR, Dykstra J, Veine C, Schuen A. Exploring the effects of a short-term, nature-based preschool experience: a mixed-methods investigation. Journal of Outdoor and Environmental Education. 2023.
28. Shleldrake R, Amos R, Reiss MJ. Children and Nature: A research evaluation for the Wildlife Trusts. 2019.
29. Sprague N, Berrigan D, Ekenga CC. An Analysis of the Educational and Health-Related Benefits of Nature-Based Environmental Education in Low-Income Black and Hispanic Children. Health equity. 2020;4(1):198-210.
30. Sprague NL, Ekenga CC. The impact of nature-based education on health-related quality of life among low-income youth: results from an intervention study. Journal of Public Health. 2021;44(2):394-401.
31. Taylor AF, Butts-Wilmsmeyer C. Self-regulation gains in kindergarten related to frequency of green schoolyard use. Journal of Environmental Psychology. 2020;70:101440.
32. van den Berg AE, Hovinga D, Joven M, Steensma R, Maas J. Strengthening the pedagogical use of the outdoor area at nature-based daycare centers: An intervention study. Urban Forestry & Urban Greening. 2024;92:N.PAG-N.PAG.
33. van Dijk-Wesselius JE, Maas J, Hovinga D, van Vugt M, van den Berg AE. The impact of greening schoolyards on the appreciation, and physical, cognitive and social-emotional well-being of schoolchildren: A prospective intervention study. Landscape and Urban Planning. 2018;180:15-26.
34. Waliczek TM, Bradley JC, Zajicek JM. The Effect of School Gardens on Children's Interpersonal Relationships and Attitudes Toward School. HortTechnology. 2001;11(3):466-8.
35. Wallner P, Kundi M, Arnberger A, Eder R, Allex B, Weitensfelder L, et al. Reloading Pupils' Batteries: Impact of Green Spaces on Cognition and Wellbeing. Int J Environ Res Public Health. 2018;15(6).
36. Whitburn J, Abrahamse W, Linklater W. Do environmental education fieldtrips strengthen children's connection to nature and promote environmental behaviour or wellbeing? Current Research in Ecological and Social Psychology Vol 5, 2023, ArtID 100163. 2023;5.
37. Wood C, Gladwell V, Barton J. A repeated measures experiment of school playing environment to increase physical activity and enhance self-esteem in UK school children. PloS one. 2014;9(9):e108701.
