## Supplemental file D Results Table for "Impacts of green space interventions in educational settings on children and young people’s mental wellbeing: a systematic review"

| | Mental wellbeing outcome(s) | Tool(s) | Statistical method(s) | Main finding | Additional finding 1 | Addition al finding 2 | Interaction effects | Summary of significant effects | Limitations | Study quality |
|---|---|---|---|---|---|---|---|---|---|---|
| Amicone (2018) | *Study 1*<br>1.Sustained and selective attention | Bells test(revised for study of attention in childhood) | Protected t-tests | When recess occurred in built environment, there was a significant improvement in Bell's test scores between T1 (M = 31.85, SE = 0.31) to T2 (M = 32.61, SE = 0.30), t(75) = 2.45; p = 0.016; d = 0.40.<br>When recess occurred in the built environment, participants did not report a significant difference between T1 (M = 31.55, SE = 0.34) and T2 (M = 31.77, SE = 0.34) in their sustained and selective attention scores, t(75) = 0.73; p = 0.47; d = 0.12. | | | | Significant positive effect | Participants performed well in both conditions; ceiling effect or learning effect may have occurred .<br><br>Activity undertaken in break time not relaxing (competitive team play) | Modera te |
| | 2.Working memory | Digit span test (from WISC-IV intelligence scale) | Protected t-tests | When experiencing their recess time in the natural environment, pupils reported a significant improvement in digit span test scores from T1 | | | | Significant positive effect of the environment on working memory | | |

| | | | | | | | | | |
|---|---|---|---|---|---|---|---|---|---|
| | | | | (M = 15.22, SE = 0.34) to T2 (M = 16.38, SE = 0.38), t(73) = 4.12; p<0.001; d = 0.68. When recess time occurred in the built environment, participants did not report a significant difference between T1 (M = 15.42, SE = 0.41) and T2 (M = 15.86, SE = 0.38) in their working memory score, t(73) = 1.55; p = 0.12; d = 0.26 | | | | | |
| | 3. Impulse control | Go-no-go test (from BIA for assessment of children with ADHD) | Protected t-tests | Results showed that in the natural environment, participants did not increase their impulse control from T1 (M = 16.85, SE = 0.43) to T2 (M = 16.79, SE = 0.42), t(75) = 0.19; p = 0.85; d = 0.03. Neither they did in the built environment, where no difference emerged between T1 (M = 16.59, SE = 0.40) and T2 (M = 16.97, SE = 0.31) in their impulse control score, t(75) = 1.04; p = 0.30; d = 0.17. | | | | No effects found | Go-no-go test may have been too easy for the sampled children, explaining the ceiling effect that might have occurred. |

| | | | | | | | | | | |
|---|---|---|---|---|---|---|---|---|---|---|
| | *Study 2* 4. Sustained and selective attention | As for study 1 but with variation in target stimulus between time 1 and time 2 | Mixed model ANOVA Series of mean difference z-tests | At T1, no significant difference in attention between natural (M = [illegible]0.08; SD = 1.21; N = 18) and built environments (M = 0.102; SD = 0.78; N = 17): z(33) = 0.54; p = 0.59, indicating that selective attention was at the same level before the manipulation occurred. Mean difference z-test for difference between NE and BE conditions at time 2: z(33) = 2.47; p = 0.007 | Interaction effect of time and condition on the attention score: F(1,33) = 10.00; p = 0.003; $\eta_p^2$ = 0.233 | | | Significant positive effect | Children in BE condition had to be instructed not to play in natural environment and this could have been seen as a limitation; attention scores decreased in the BE condition in study 2. | |
| Anabirtate (2021) | Attention restoration | Attention Network Test (ANT) | Linear mixed effects model | Exposure x time interaction effect not significant for any combination of response and school (p values ranged from 0.109 to 0.910) | | | | No evidence that participation in a relaxing activity in green versus grey spaces affected students' performance in attention tests | Study area located in green surroundings. The study sites were small and medium sized towns in the Basque country, located near the sea and mountains and therefore there might be less impact of green space | Modera te |

| | | | | | | | | | | |
|---|---|---|---|---|---|---|---|---|---|---|
| | | | | | | | | | exposure (prior studies showed more benefit in urban areas). Environment al noise from busy roads near all the test sites may have interfered with attention restoration. One hour of exposure to green space may have been insufficient. | |
| Barrable (2021) | Positive affect | Positive Affect, Negative Affect Scale for Children (PANASC) | 1. Mixed effects GLM fitted to predictor variables<br>2.Paired Wilcoxson signed rank test for individual affect scores | Results from minimal adequate regression models:<br>Small-to-medium sized effect (McFadden's pseudo-R-squared = 0.13) increase in positive affect post-activity.<br>Small sized effect of drop in negative affect (McFadden's pseudo-R-squared = 0.02) | | | | Small to medium sized effect in increase in all dimensions of positive affect ($p<0.001$).<br>Of the negative affect dimensions, only "miserable" "sad" and "angry" had a | Only 20 participants from 2 schools able to complete follow-up questionnaire 8 weeks after intervention. Changes in affect could have been caused by a visit to a nature | Weak |

| | | | | | | | | | | |
|---|---|---|---|---|---|---|---|---|---|---|
| | | | | | | | | significant change (P<0.05). | reserve per se rather than the mindfulness intervention in nature. No control group | |
| Barton (2015) | Self-esteem (SE) | Modified 10-item Rosenberg Self-Esteem Scale | Two-way ANCOVA to examine effect of intervention type and school location on change in self-esteem | Two-way ANCOVA showed no significant main effects on change in SE due to type of intervention (p>0.05) or school location (rural or urban) (p>0.05) | | | Two-way ANCOVA showed no significant main effects on change in SE due to interaction between type of intervention and school location (p>0.05) | No significant main effects on change in self-esteem found for type of intervention, school location or interaction between these. | Duration of intervention (1 week) may have been insufficient to produce significant effect. Order of interventions not randomised. | Weak |
| Bates (2018) | Perceptions of safety | Teacher and caregiver surveys; Likert scale of 5 choices; retrospective comparison with behaviour pre-renovation | Independent samples t-tests | Caregivers' survey (scale -2 to +2) at T1 = 0.77, T3 = 1.03; t-value -1.39; p > 0.05 | Teachers' survey: at T1 = 1.24, T3 = 1.21; t-value 0.15; p > 0.05 | | | Caregivers and teachers retrospectively reported that compared to pre-renovation schoolyards appeared safer, students | Schoolyards were only assessed post-renovation. Co-intervention was additional renovation of non-green play spaces. | Weak |

| | | | | | | | | | |
|---|---|---|---|---|---|---|---|---|---|
| | | | | | | | | experienced fewer injuries, and less bullying and gang-related activity. No significant changes in these over time, post-renovation, over 1-year period. | No student self-report data. Positive perceptions by teachers and caregivers may have been impacted by community-engaged planning process. |
| | Reported injuries | | | Caregivers' survey (scale -2 to +2) at T1 = 0.80, at T3 = 0.90; t-value -.058; p > 0.05 | Teachers' survey at T1 = 0.77, at T3 = 0.69; t-value 0.37; p > 0.05 | | | | |
| | Reported bullying | | | Caregivers' survey (scale -2 to +2) at T1 = 0.66, at T3 = 0.65; t-value 0.07; p > 0.05 | Teachers' survey at T1 = 0.53, at T3 = 0.53; t-value 0.03; p > 0.05 | | | | |
| | Reported gang-related activity | | | Caregivers' survey (scale -2 to +2) at T1 = 0.68, at T3 = 0.86; t-value -1.07; p > 0.05 | Teachers' survey at T1 = 0.57; at T3 = 0.77; t-value - | | | | |

| | | | | | | | | | | |
|---|---|---|---|---|---|---|---|---|---|---|
| | | | | | 0.83; p > 0.05 | | | | | |
| | Student interactions | Behavioural mapping using SOCARP codes | Chi-square analyses | Changes in observed social interactions over time, from T1 to T2 (6 months later): $\chi^2 = 98.80$, $p < 0.001$ | Proportion of negative interactions stable at T2 compared with T1 | Greater percentage of children interacting with each other socially at T2. | | Social interactions showed statistically significant increase over a 6-month period post-schoolyard renovation | | |
| Block 2012 | Wellbeing | KIDSCREEN-10 (quality-of -life score) | Random effects linear regression models Generalized estimating equations | Statistic for difference in mean quality-of-life scores between programme and comparison schools = 1.23(SD 0.7); CI -0.2 to 2.7; p = 0.09 | | | | Weak evidence of a positive programme effect on quality-of-life scores, but not reaching statistical significance at 5% level. | There may have been a "ceiling effect" on child cooperative behaviour scores as high scores recorded at baseline left little room for improvement. 1-year period between baseline and follow-up evaluation may not have been long enough for any changes to | Moderate |

| | | | | | | | | | |
|---|---|---|---|---|---|---|---|---|---|
| | | | | | | | | | cooperative behaviour to be extended beyond the garden classes. Sample size on teacher questionnaires was too small to yield significant finding. KIDSCREEN-10 may not be sensitive to change. |
| | Cooperative behaviours | Modified section of Robinson and Zajicek's 2005 scale for self-, parent- and teacher-reported cooperative behaviours | | Adjusted test statistic for self-reported child cooperative behaviour scores = 0.31; 95% CI -0.16 to 0.77; p = 0.2 | Adjusted test statistic for parent-reported child cooperative behaviour scores = 0.11 (SD 0.2); 95% CI -0.56 to 0.35; p = 0.7 | Adjusted test statistic for teacher -reported child cooperative behaviour scores = -0.42 (SD 0.5); 95% CI -1.4 to 0.57; p= 0.4 | | No evidence of true difference in child cooperative behaviour scores between programme and comparison groups | |
| | Student engagement | Teacher perceptions; | | Adjusted test statistic for "Teacher strongly agrees student looks | Unadjusted test statistic for | Adjusted test statistic | | No evidence of true difference | |

| | | | | | | | | | | |
|---|---|---|---|---|---|---|---|---|---|---|
| | and social behaviour | survey; 4-point scale | | forward to coming to school” = 0.57; CI 0.24-1.36; p = 0.2 | “Teacher strongly agrees student social behaviour in this school is good” = 0.82; 95% CI 0.17 to 1.50; p = 0.2 | for “Teacher strongly agrees students cooperate well with other students in the school” = 0.51; 95% CI 0.13 to 2.05; p = 0.3 | | between programme and comparison schools in teachers’ perceptions of student engagement and social behaviour | | |
| Brussoni (2017) | Conduct | Strengths and Difficulties Questionnaire (SDQ) | Wilcoxon signed-rank test | Change in DSQ peer problems scale: Median T1 = 2.3, T2 = 2.0; z = -2.10, p = 0.036. | | | | Significant decrease on peer problems scale from T1 to T2 (pre-to post-intervention) | Difficulty isolating intervention effects from typical child development. Wellbeing and play potentially affected by season during data collection period (T1=winter, T2=spring). | Moderate |
| | Social behaviour | Preschool Social Behaviour Scale-Teacher Form (PSBS-T) | | PSBS depressed affect score change: Median T1 = 6.0, T2 = 3.0; z = -2.24, p = 0.03 | | | | Significant decrease in depressed affect score from T1 to T2 | | |
| | | Play observations (coded)-measuring | Generalised linear mixed effects | OR (CI) for prosocial behaviour increase from T1 to T2 = 2.81(1.17-6.91) in | OR (CI) for antisocial behaviour increase in centre A | | | Prosocial behaviour increased significantly in centre A and | | |

| | | | | | | | | | | |
|---|---|---|---|---|---|---|---|---|---|---|
| | | prosocial behaviour | models (GLMM) | Centre A, 0.17 (0.05-0.63) in Centre B | 1.40(0.47-1.43), in centre B 0.16(0.03-0.75) | | | decreased in centre B. Antisocial behaviour decreased in both centres, but the decrease was less in centre A | | |
| Carrus (2015) | Quality of affective state | Observed frequency of positive and negative affect-related behaviours | ANOVAs | Positive affect only increased in frequency on garden days **after** the free play activity in nature ($F(1, 18) = 9.14$, $p = .007$, partial $h^2 = 0.34$) | | | | Significant improvement in affect on garden days after free play activity in nature | Inter-rater reliability not assessed. Single observer per child. Observers may have been aware of hypotheses. Lack of control group. Positive effects may have been caused by simply being outside classroom. | Weak |
| | Quality of social behaviour | Frequency of positive and negative | ANOVAs | More frequent positive relational behaviour on garden days : ($M = 0.12$; $SD =$ | | | | Free play activity in nature associated | | |

| | | | | | | | | | | |
|---|---|---|---|---|---|---|---|---|---|---|
| | | relational behaviours | | 0.012) compared to NGDs (M = 0.05; SD = 0.06), but only in observations made after children were exposed to nature, after the free-play activity (F(1, 18) = 4.99, p = .038, partial η2 = .22). | | | | with significant increase in positive relational behaviour | | |
| Dopko (2019) | Mood | Shortened mood measure from Positive and Negative Affect Schedule for Children | Paired samples t-tests | Positive affect $d_z$(95%CI) = 0.28 (0.03-0.54) p = 0.03<br>Negative affect $d_z$ = 0.44 (0.18-0.71) p = 0.001<br>Vitality $d_z$ = 0.37 (-0.14 to 0.36) p = 0.37<br>Pleasant affect $d_z$ = 0.13 (-0.12 to 0.39) p = 0.31<br>Fun today $d_z$ = 0.21 (-0.05 to 0.46) p = 0.12 | | | | Significantly more positive and negative affect reported at nature school vs museum. No significant difference in vitality, low arousal pleasant affect, or fun had. | There were differences other than nature immersion between the two locations (less structure, more autonomy in nature school). Children may have experienced difficulties understanding the tangram and windfall tasks. There was a set order of locations (nature | Weak |

| | | | | | | | | | |
|---|---|---|---|---|---|---|---|---|---|
| | | | | | | | | | school first) so some learning may have occurred on the tasks. Visiting gift shop at museum may have primed responses. |
| | Pro-sociality | Windfall task | | Buying stuff $d_z$ = 0.09 (-0.22 to 0.40) p = 0.56<br>Charity $d_z$ = 0.32 (0.01-0.63) p = 0.047<br>Gifts $d_z$ = -0.31 (-0.63 to 0.002) p = 0.051<br>Save $d_z$ = -0.07 (-0.38 to 0.24) p = 0.66 | | | | Significantly more money donated to charity in nature task vs museum. Amount of money allocated for gifts was marginally significant: less money for gifts was allocated at nature school. No significant difference on buying stuff for themselves and saving between two settings. | |

| | | | | | | | | | | |
|---|---|---|---|---|---|---|---|---|---|---|
| | | Tangram task | | Easy $d_z$ = 0.32 (0.01-0.63) p = 0.046 Medium $d_z$ = 0.30 (0.01 to 0.61) p = 0.06 Hard $d_z$ = -0.46 (-0.78 to -0.14) p = 0.005 | | | | At nature school significantly fewer hard tangrams and larger number easy tangrams assigned to others (suggests higher pro-sociality). | | |
| Ernst (2019(a)) and (2019(b)) | Executive function skills (cognitive flexibility, inhibitory control, working memory) | Minnesota Executive Function Scale (MEFS) | ANOVA | No significant difference between the nature and non-nature participants, when controlling for age, gender, prior participation, and pretest level (F(1) = 0.28, p = 0.60 | | | | No significant difference in outcomes between children attending nature based and non-nature-based preschools | Both groups had increases in executive function skills scores that were higher than expected-the authors thought this was likely to be due to attendance at a play based preschool and that both an indoor and outdoor environment were beneficial. | Weak (a); Weak (b) |
| | Resilience (initiative, self- | Devereux Early Childhood | Repeated measures ANOVAs | Total protective factors score: | Initiative score: | Self-regulation score: | | For nature preschoolers there was | Small sample size of comparison | |

| | | | | | | | | | | |
|---|---|---|---|---|---|---|---|---|---|---|
| | regulation, attachment) | Assessment for Preschoolers ($2^{nd}$ edn) (DECA-P2)- 27 items on subscales of initiative, self-regulation and attachment | | Growth in teacher ratings for nature preschoolers $F(1,76) = 16.32$, $p < 0.001$ Parent rating $F(1,76) = 7.13$, $p = 0.009$. | Growth in teacher rating for nature preschoolers $F(1,76) = 32.48$, $p < 0.001$ Parent rating $F(1,76) = 13.58$, $p < 0.001$ Non-nature preschool growth in teacher ratings $F(1,10) = 30.63$, $p < 0.001$ | Growth in teacher rating for nature preschoolers $F(1,76) = 10.56$, $p = 0.002$ Parent rating $F(1,76) = 10.34$, $p = 0.002$ | | significant growth in parent and teacher ratings on initiative and self-regulation and in the total protective score. For non-nature preschoolers the only significant growth was in teacher ratings of initiative. | group-the authors comment this group served as comparison group rather than true control group. Homogenous nature of participants (primarily Caucasian and middle to high socio-economic status) | |
| Harvey (2020) | Wellbeing | KISDCREEN-27 (results not disaggregated by domain) | Wilcoxon signed rank tests | Mean difference in wellbeing score change over time between programme and control groups = 6.87 (CI 1.01-12.74) $Z = 2.86$, $p = 0.004$ | | | | Children in intervention group increased scores significantly more than those in control | Numbers in control not matched to experimental programme Social desirability bias Common method bias (reliance on introspective measures) | Moderate |
| | Mood | Positive and Negative Affect Schedule for | Positive mood change across course of | Improvement in mood between start and end of each nature session ($F(1,2809) = 72.49$; $p < 0.001$; not affected by | | | | Significant increase in mood scores over course of nature | | |

| | | | | | | | | | | |
|---|---|---|---|---|---|---|---|---|---|---|
| | | Children (PANAS-C) | nature sessions $Z = 6.43$; $p < 0.01$ | session number (no interaction, $p = 0.39$) | | | | sessions. No comparison with control. | | |
| Kelz (2015) | Wellbeing | Basler Wellbeing Questionnaire (intra-psychic balance sub-scale) | Planned Comparisons Method | Comparison of mean wellbeing scores at experimental school post-intervention with pre-intervention and control scores $F(1,190.3) = 7.63$, $p = 0.006$, $d = 0.31$ | | | | Significant difference in intra-psychic subscale scores after installation of new schoolyard | Concurrent introduction of additional sports equipment and seating might have acted as co-intervention. Experimental and control schools different school types. First and second measurements carried out in different seasons. Renovated schoolyard "weak" effect as financial constraints caused limitations to landscaping. | Weak |
| | | Recovery-Stress Questionnaire (R-SQ) | | $F(1,172.3) = 3.78$, $p = 0.053$, $d = 0.18$ | | | | Marginally statistically significant difference in recovery from stress | | |
| Largo-Wight (2019) | Child happiness and wellbeing | Children's "Face Scale" Survey (3-point scale) | Independent samples t-test | For difference in response between conditions, $t = 1.71$, $p = 0.097$ | | | | No significant effect of condition on self-reported | Small sample size (two teachers, 36 children). | Moderate |

| | | | | | | | | | | |
|---|---|---|---|---|---|---|---|---|---|---|
| | | rating on one happiness question | | | | | | child happiness | Possible participant fatigue over time (6 weeks) when using the Faces Survey as was used daily. | |
| | | | Mixed effect model | For effect of environment on smiley face percentage F = 0.6489, p = 0.4256 | | | | | | |
| | Child behaviour | Count of teacher redirections per minute/child | Independent samples t-test | Difference in means between conditions (indoor and outdoor), average for both teachers = 0.0127, t = 1.487, p = 0.1383 | Difference in means between classrooms for more experienced teacher = 0.0254, t = 2.27, p = 0.025 | | | There was a reduction in teacher redirects in the outdoor condition; this was only statistically significant for one of the teachers | | |
| | Child attention and focus | Count of students off task per minute | Independent samples t-test | Difference in means between conditions, for both teachers, t = 1.17, p = 0.244 | | | | Nature did not have a statistically significant effect | | |
| Mason (2022) | Affective state | Positive and Negative Affect Schedule (PANAS) (shortened version) | Linear mixed models | For the main effect of green environment on positive affect, no significant difference was found pre- and post-lesson in greenness (B = 0.54 (CI -0.36-1.43), p = 0.240). See adjacent findings for the role of emotional difficulties as a moderator. | | Positive affect of students with lower emotional difficulties did not vary as a function of the environ | Students with higher emotional difficulties reported lower negative affect after the lesson in the green than after the lesson in the | For students with lower emotional difficulties, negative affect reported after the lesson did not vary as a function of the environment in which the | Emotional problems were the only possible moderator examined between the environment and outcome variables. A larger sample would allow other | Weak |

| | | | | | | | | | |
|---|---|---|---|---|---|---|---|---|---|
| | | | | Greenness did not have an effect on negative affect either ( B = -0.05 (CI -0.15-0.96), p = 0.399). Further findings describe interaction between time, environment and emotional difficulties (measured for each student prior to intervention) Students with higher emotional difficulties reported statistically significant increase of positive affect after the school lesson in greenness than after the lesson in the classroom (B = 0.97, SE = 0.37, t = 0.64, p = 0.01) | | ment where the lesson took place (B = -0.01, SE = 0.37, t = -0.04, p = 0.97) | classroom, however the difference between the 2 environments did not reach statistical significance ( B = -0.08, SE = 0.04, t = -1.71, p = 0.09) | lesson took place ( B = -0.02, SE = 0.04, t = -0.48, p = 0.63) | individual differences to be examined. The emotional difficulty scale used had doubtful reliability. Equivalence of the lessons given in the different environments was not assessed. |
| | Selective and sustained attention | Bells test | | For selective attention main effect of environment, B=2.79 (P<0.014)<br>For sustained attention, B=0.71 (P < 0.029) | | | | Children had higher selective and sustained attention scores after the green lesson. Both children with higher and lower emotional difficulties had | |

| | | | | | | | | | | |
|---|---|---|---|---|---|---|---|---|---|---|
| | | | | | | | | better scores in the greenness than indoor | | |
| Montgomery (2022) | Wellbeing | Kidscreen-27 (results not disaggregated) | Mann-Whitney-Wilcoxon tests Spearman rank correlation Generalised linear mixed models | Overall, no significant change in wellbeing scores ($\chi^2$ = 2.8, df = 3, p > 0.05) after the activities with the intervention and control groups. | Children in intervention group with initially low scores (below 100, n = 46) for wellbeing had significant improvements in wellbeing (before = 92.2., after = 103.8, V = 80.0, p < 0.001). | Due to low numbers in the control group of those with initially low wellbeing (n = 18) an analysis of a moderating effect of initially low wellbeing was not possible. | | The main hypothesis that children who participated in wellbeing programmes experience an increase in wellbeing was not supported, however children with initial low wellbeing scores did experience a significant increase | Children's biodiversity perceptions were assessed using drawings-as result some children may not have completed the task fully as ran out of time. | Weak |
| | Resilience | Child and Youth Resilience Measure 12 (CYRM-12) | Generalised linear mixed models Tukey post-hoc tests Wilcoxon signed-rank tests | No significant change in the resilience scores after the activities within the intervention and control groups ($x^2$=0.01, df=3, p>0.05) | Children in intervention group with initially low wellbeing and resilience scores had significant improvem | | | Children who participated in the intervention did not experience a significant change in resilience, however children in the intervention | | |

| | | | | | | | | | | |
|---|---|---|---|---|---|---|---|---|---|---|
| | | | | | ents in resilience (before=23, after = 27), V = 36.5, p < 0.01). | | | group with initially low wellbeing and resilience scores had significant improvements in resilience. | | |
| Moula (2023) | Wellbeing | Personal Wellbeing Index-School Children (PWI-SC) (not disaggregated by domain) | Paired-sample Sign test | Small effect size P = 0.28, Cohen's d = 0.35 for change in wellbeing score<br>56 out of 95 children rated their subjective wellbeing higher post-intervention | | | | No statistically significant difference was observed | There was no control group. | Weak |
| Mueller (2017) | Directed attention | Continuous Performance Test (CPT) | ANCOVA | F(1,68) = 0.44, p = 0.51, $\eta^2 = 0.01$ | | | | No significant differences for cognitive skills | Lack of randomisation of individuals. Large amount of nature exposure outside school as district in forested area. Short-term follow-up. Parents and teachers not blinded to the intervention. | Weak |
| | Inhibition | Heads-shoulders-knees-toes task (HSKT) | | F91,83) = 0.92, p = 0.76, $\eta^2 = 0.00$ | | | | | | |
| | Social skills and psychological health | Social skills rating scale (SSRS) | | Teacher Report<br>Assertiveness F(1,80) = 40.44, p<0.01, $\eta^2 = 0.34$<br><br>Cooperation F(1,81) = 20.20, p<0.01, $\eta^2 = 0.20$<br><br>Self-control F(1,79) = 37.56, p < 0.01, $\eta^2 = 0.32$ | Parent Report<br>Assertiveness F(1,46) = 6.56, p = 0.01, $\eta^2 = 0.13$<br>Social responsibility F(1,44) = 5.39, p = 0.03, $\eta^2 = 0.11$ | | | Large effect sizes and significant differences for the nature kindergarten in the teacher report on social skills. Parent reports on assertiveness and social | | |

| | | | | | | | | | | |
|---|---|---|---|---|---|---|---|---|---|---|
| | | | | Externalizing behaviour $F(1,79) = 2.58$, $P = 0.11$, $\eta^2 = 0.03$<br><br>Internalizing Behaviour $F(1,81) = 4.35$, $p=0.04$, $\eta^2 = 0.05$ | Cooperation $F(1,48) = 3.88$, $p = 0.06$, $\eta^2 = 0.08$<br>Self-control $F(1,48) = 1.14$, $p = 0.29$, $\eta^2$ 0.02<br>Externalizing Behaviour $F(1,49) = 1.63$, $P = 0.25$, $\eta^2 = 0.03$<br>Internalizing Behaviour $F(1,51) = 0.17$, $p = 0.68$, $\eta^2 = 0.00$ | | | responsibility also significantly higher for nature kindergarten. | | |
| Mygind (2009) | Social relations | Questionnaire with 4-point Likert scale | Wilcoxon Signed Ranks Test | Sum score for 10 social relations statements showed significant difference in means between forest and classroom settings $p < 0.001$ | | | | Social relations measures were significantly higher in the forest setting when | Small sample size, asymmetrical gender distribution. | Weak |

| | | | | | | | | | | |
|---|---|---|---|---|---|---|---|---|---|---|
| | | | | | | | | analysed as a sum score. | | |
| Pirchio (2021) | Psycho-physical wellbeing (results not disaggregated) | 5 items on 4-step Likert scale taken from WHO-5 | ANOVA | | | | **Study 1:** Effect of contact with nature on psycho-physical wellbeing for intervention group, as a two-way interaction effect of group (intervention vs. control) by time (pre-post) [ $F_{(318,1)}$ = 16.7; p = 0.000 ] **Study 2**: [ $F_{(319,1)}$ = 24.428; p = 0.000] | **Studies 1 and 2**: Significant effect of contact with nature on psycho-physical wellbeing but not on life satisfaction | Timing and intensity of interventions may have impacted on absence of significant findings in relation to life satisfaction (and other outcomes not included here). | Weak |
| | Life satisfaction | Seven items on satisfaction with school, living environment and school | | | | | **Study 1**: No significant interaction effect [ $F_{(306,1)}$ = 0.001; p = 0.961 ] | | | |

| | | | | | | | | | | |
|---|---|---|---|---|---|---|---|---|---|---|
| | | on 4-step Likert scale | | | | | **Study 2**: [ $F_{(310,1)}$ = 0.636; p = 0.426] | | | |
| Pollin (2021) | Emotions | Emotion diary | Mann-Whitney U-test | 3 out of 10 emotions were perceived significantly (p < 0.001) more often (happiness, pride, surprise/wonder) in the school garden and 2 significantly (p , 0.05) more often in the classroom (disgust, fear/anxiety) | | | | Significant differences in perception of 3 positive emotions (happiness, pride, surprise/wonder) in school garden and 2 negative emotions (disgust, fear/anxiety) in classroom. | School garden environment and classroom environment differed in teachers personal and social role during lessons as well as organisation of lessons and proportion of theory and practice. Unclear validity of the emotion diary. Memory errors due to delay in recording emotions 60 minutes after activities. | Weak |

| | | | | | | | | | | |
|---|---|---|---|---|---|---|---|---|---|---|
| Raney (2019) | Social interactions | System for Observing Children's Activity and Relationships During Play (SOCARP) | Linear Mixed Models | Student physical and verbal conflict rates decreased below pre-greening rates after 4 months for the experimental group (p<0.001).<br><br>Playground greening resulted in a significant decrease in minutes spent alone (mean difference = -2.22, 95% CI = -1.7, -2.7, p<0.001) and a significant increase in the number of minutes spent in small groups (mean difference = 1.7, 95% CI = 0.9, 2.6, P <0.01), which was positively correlated with the amount of time spent in small groups (r=0.26, p<0.001) | | | | Significant decrease in conflict rates in experimental group, as well as decrease in time spent alone | Small size of control group. Only one experimental site. | Moderate |
| Raney (2021) | Social interactions | System for Observing Children's Activity and Relationships During Play (SOCARP) | Linear Mixed Models | Study found interaction effect between condition and study phase for the number of prosocial (p = 0.03) and antisocial interactions (p = 0.003). Prosocial interactions were | | | | | Note: This is a continuation of the study reported in Raney (2019), with outcomes measured at 16 months post-greening | Moderate |

| | | | | | | | | | | |
|---|---|---|---|---|---|---|---|---|---|---|
| | | | | more frequent, and antisocial interactions less frequent, at the experimental site at 16 months. No differences in pro-or antisocial interactions were observed at the control locations over time. | | | | | Findings may not be generalisable to other schools (in terms of poorer adaptation to climate changes throughout the year and fewer sports facilities, which were present in both the control and experimental schools).Additional factors that influence recess behaviour (e.g. instructional use of schoolyards by teachers) were not recorded. | |
| Reed (2013) | Self-esteem | Rosenberg Self Esteem Scale (RSE) | Two-way repeated measures analysis of | No main effect for exercise condition ( $F(1,74) = 0.02$, $p = 0.898$ ) and no | RM t-tests showed there were no | | | Green exercise did not create additional | RSE typically used as trait scale | Moderate |

| | | | | | | | | | | |
|---|---|---|---|---|---|---|---|---|---|---|
| | | | variance (rm-ANOVA) Repeated measures t-tests | interaction between exercise and terrain condition. F(1,74) = 0.13, p = 0.72 | significant differences between green and control exercise in terms of self-esteem (t(75) = 0.13, p = 0.72) | | | improvement in self-esteem above that observed in control condition. | Control condition artificial; contaminated by researcher presence. Terrain differences between conditions (green condition harder) | |
| | Enjoyment | Visual analogue scale (VAS) | Repeated measures t-tests | RM t-tests showed there were no significant differences between green and control exercise in terms of enjoyment (t(75) = 0.43, p = 0.66) | | | | Perceived enjoyment was similar in green and control conditions. | | |
| Roberts (2017) | Wellbeing | Warwick-Edinburgh Mental Wellbeing Scale (WEMWBS) | Independent sample t-tests Repeated measures analysis of variance (ANOVA) Paired sample t-tests | WEMWBS (time 2 versus time 1) t = -1.43, df122, p = 0.078, Cohen's d = 0.12<br><br>For males only, p = 0.025<br>Females only p = 0.463 | Effect of time (times 1, 2 and 3) on WEMWBS scores: F(1.78, 217.07) = 4.75, p = 0.01 | Males maintained significantly higher levels of WEMWBS (*t*(33), = -2.94, *p* = .003) at time 3 (WEMWBS: M = 3.87, SD = 0.56; compare | | No significant difference in outcomes between students who had completed forest school and those in control group pre- and post-forest school experience. When analysed by separate | Limited generalisability as only one school enrolled. Sensitivity to change of instruments used may be limited, and exposure time (5 weeks) insufficient to produce | Moderate |

| | | | | | | | | | |
|---|---|---|---|---|---|---|---|---|---|
| | | | | | | d with time 1 (WEMWBS: M = 3.56, SD = 0.66. | | gender, significant difference over time in males only. Significant effect at T3 (3 months post completion of forest school) | detectable change. Social desirability bias. Control group did not include non-nature aspects of the intervention (fun activities outside the classroom). |
| | Resilience | Sense of Mastery Scale (SoM), subscale of Resiliency Scales for Children and Adolescents (RSCA) | | SoM (time 2 versus time 1) t = -2.19, df122, p = 0.015, d = 0.19<br><br>For males only, p = 0.029<br>For females only, p = 0.130 | Effect of time (T1, T2 and T3) on SoM scores: F (1.71, 208.50) = 13.07, p <0.001) | Females maintained significantly higher levels of SoM at time 3 (*t*(20), = -2.61, *p* = .009; M = 3.84, SD = 0.75) compared with time 1 (M = 3.49, SD = 0.53). | | Resilience, measured by SoM scale, increased significantly from time 1 to time 2 for the intervention group. When analysed by gender, significant difference over time in males only. Significant effect at T3. | |

| | | | | | | | | | | |
|---|---|---|---|---|---|---|---|---|---|---|
| Roe (2011) | Mood | 14-item version of UWIST Mood Adjective Shortlist (MACL) | ANOVA | Effect of setting on energy (EA) : F = 9.65, df = 1, p = 0.007<br>Stress (TA): F = 4.44. df = , p = 0.052<br>Hedonic tone (HT) F= 9.99, df = 1, p = 0.007<br>Anger: F = 6.665, df = 1, p = 0.020 | Difference between the good and poor behaviour groups:<br>For EA, F = 9.48, df = 1, p = 0.008<br>For TA, F = 5.45, df = 1, p = 0.034<br>For HT, F = 4.49, df = 1, p = 0.052<br>For anger, no significant difference between the two behaviour groups. | | | Significant main effect of setting on energy; borderline significant main effect on stress, significant main effect on hedonic tone and on anger. There was a significant difference between the two behaviour groups for energy and stress, and a borderline difference for hedonic tone, with the poor behaviour group benefitting most from the forest school experience. | Small sample size (n=18). Inability to control for social context between settings. Activity in school different from the forest activity. | Weak |
| Scogin (2023) | Social-emotional outcomes | Teaching Strategies GOLD (TS Gold)-focus on social-emotional | Wilcoxon signed rank test | Z values for “Manages Feelings” 2.38 (p = 0.018); “Follows Limits and Expectations” 4.11 (<0.001); “Takes Care of Own Needs” 3.21 (p = 0.001); “Balances Needs of Self and Others 0.49 (p = 0.626); “Solves Social Problems” (p<0.001); Attends and Engages 3.90 (p<0.001); Solves | | | | Statistically significant, positive growth in all social/emotional areas | Small sample<br>No control or comparison groups<br>Selective recruitment | Weak |

| | | | | | | | | | | |
|---|---|---|---|---|---|---|---|---|---|---|
| | | and cognitive development | | Problems 3.97 (p<0.001); Shows Curiosity and Motivation 2.61 (0.009) | | | | tested, with the exception of "Balances Needs" | may have occurred (children whose families were already more connected to nature may have been more likely to sign them up) | |
| Sheldrake (2019) | Wellbeing | Questionnaire with 5-point Likert scale (results not disaggregated) | Repeated measures ANOVA | Main effect of time: F(1,355) = 10.6, p < 0.001 | | | Significant interaction effect between baseline wellbeing and time. Children with low initial wellbeing showed increases in wellbeing means from 3.61 to 3.90 (p , 0.001); children with high initial wellbeing showed no significant change. | Significant increases over time overall in children's subjective wellbeing. Children with low initial wellbeing experienced greater positive change in wellbeing. | No comparison group. Heterogeneous interventions. Ceiling effect possible on some measurements. | Weak |

| | | | | | | | | | | |
|---|---|---|---|---|---|---|---|---|---|---|
| Sprague (2020) | Health Related Quality of Life | HRQoL domains (results disaggregated) | Independent *t*-tests Linear regression models | Emotional health functioning mean pre-intervention score 2.4 (SD1.2), post intervention score 4.1 (SD 0.9). p-value for difference < 0.001<br><br>Social functioning pre-intervention 2.6 (1.2), post-intervention 4.1 (0.1), p < 0.05<br><br>Overall HRQoL pre-intervention 13.5 (3.7), post-intervention 19.9 (2.8), p<0.05 | | | No significant interaction between gender and duration with change in means.<br>For age, interactions significant for social functioning and overall HRQoL (older children had greater improvements) | Significant improvement in all HRQoL domain (including social functioning, emotional health functioning and overall HRQoL) mean and median scores from pre-intervention to post-intervention | Modest sample size and duration<br>Reduced generalisability as children from urban public school system in Missouri, USA<br>No control group | Weak |
| Sprague (2021) | Health Related Quality of Life | 5 HRQoL domains including emotional health functioning (results disaggregated by domain) | *t*-tests<br>$\chi^2$ tests<br>Random effects analysis of covariance | After 15 weeks, intervention group showed statistically significant improvements in each HRQoL score (p < 0.001), whereas control group had significant reductions. | For emotional health functioning, the intervention group had significantly higher scores than the control group (+1.8) | | | Compared with control group, nature-based education group reported significantly higher HRQoL, including on the emotional health functioning score. | Relatively short duration (15 weeks)<br>Participants not randomised<br>Participants drawn from single district school system<br>Study could not account for nature experiences | Weak |

| | | | | | | | | | | |
|---|---|---|---|---|---|---|---|---|---|---|
| | | | | | | | | | outside school Questionnaire replies may have been subject to social desirability bias. | |
| Taylor (2020) | Behavioural Self-Regulation | Child Behaviour Rating Scale (CBRS) | Paired t-test ANCOVA | Significant improvement from pre to post ($p < 0.001$) Except girls in winter/spring semester ($p = 0.4676$) | The effect of total time outdoors was highly significant ($p = 0.01$) in the winter/spring model. In this model, the interaction between gender and total time outdoors was non-significant ($p = 0.97$). | In the autumn CBRS model total time outdoors did not significantly predict CBRS scores. | In the winter CBRS data, a significant interaction existed between frequency of greenspace curriculum and gender. Girls in the high frequency classes performed significantly better than girls in the low frequency classes (mean adjusted difference = 2.3, 95% CI | While the frequency of the greenspace curriculum sessions in the winter/spring study did not make a significant difference for boys, it had a significant impact on girls' CBRS scores. While frequency of the greenspace curriculum did not make a difference in boys' HTKS scores, it did have a significant | Effect sizes (partial eta squared) small (0.01-0.03). Intervention periods limited-nine weeks and twelve weeks. Not clear from data whether benefit to girls would level off over time. CBRS measure susceptible to bias as raters not blinded to intervention. HTKS measure has | Strong |

| | | | | | | | | | |
|---|---|---|---|---|---|---|---|---|---|
| | | | | | | | = 0.4 to 4.3). Girls in the low frequency classes performed worse than boys in the low frequency classes (mean adjusted difference = 3, 95% CI = 0.7 to 5.3). | effect on girls' HTKS scores. More total time weekly in greenspace was also related to higher self-regulation scores, but this relationship was only detected with the winter/spring CBRS, for both boys and girls, and autumn HTKS, just girls. | been shown to have floor effects in some children with low scores. Significant differences between the winter/spring and autumn study periods in terms of sample size, time of year, schoolyard characteristic s. |
| | | Head-Toes-Knees-Shoulders Task (HTKS) | | Significant improvement from pre to post ($p < 0.001$) | In the autumn HTKS model the response to total time spent outside weekly differs between boys and girls. Boys performed similarly for all possible quantities of time spent | In the autumn term girls in the high frequenc y group scored significan tly higher than low frequenc y girls (6.0-point differenc e, CI 1.7-10.3) and significan tly higher | | | |

| | | | | | | | | | | |
|---|---|---|---|---|---|---|---|---|---|---|
| | | | | | outside, whereas girls who spent more time outside tended to score higher on the HTKS post-intervention (p = 0.01). | than high frequency boys (7.4-point difference, CI 3.4-11.5) | | | | |
| Van den Berg (2024) | Wellbeing (emotional) | SICS (ZiKo) instrument 5-point wellbeing scale | ANCOVA | Mean score in outdoor area = 3.54, SE = 0.10, in control locations M = 3.25, SE = 0.80. After controlling for age, gender and location size this was a significant difference. F (1,106) = 5.67, p = 0.02, $\eta^2_p$ = 0.05 | | | Difference in wellbeing between control and intervention locations mostly applies to boys, as indicated by a marginally significant interaction effect, *F* (1,105) = 3.19, *p* = .08, *Ηp2* = .03. | | Large sample; objective measures and control groups. Tested impact of active engagement with nature (by considering effect of caregivers' pedagogical skills). Control and intervention locations not well matched. Post design | Moderate |
| | Involvement | SICS (ZiKO) instrument | ANCOVA | Children at intervention locations were about equally | | | Differences between interventio | | | |

| | | | | | | | | | | |
|---|---|---|---|---|---|---|---|---|---|---|
| | | 5-point involvement scale | | involved during outdoor play, M = 3.30, SE = 0.11, as children at control locations, M + 3.32, SE + 0.14 | | | n and control locations are moderated by gender (P = 0.008) with boys at intervention locations showing more involvement than boys at control locations, and the opposite being the case for girls. | | only, without measurement at control and intervention locations at baseline. Interobserver reliabilities not calculated as observers compared observations for practical reasons. Also, not all visits to the childcare centres could be scheduled at same interval past the completion of the intervention period. Data collection was limited to a single day at each centre. | |
| | Social behaviour | Observed time playing or spending time alone or with others | | No differences found in social behaviour between intervention and control locations, neither as main effect, nor in interaction with age or gender (Ps > 0.42) | | | | | | |
| Van Dijk-Wesselius (2018) | Emotional functioning | Subscale emotional functioning | | No significant difference in means between control and intervention groups | | | Gender and group did not moderate | No evidence for significant impact of greening | Randomisation of schools not possible so selection | Weak |

| | | | | | | | | | |
|---|---|---|---|---|---|---|---|---|---|
| | | on paediatric QoL scale | | No significant interactions between time and condition, $p > 0.41$. | | | the results, $p > 0.30$. | schoolyards on children's emotional functioning. | bias could have occurred. Results might not be generalisable to rural schools. Actual greening of schoolyards was modest in some cases and may have led to underestimation of effect of greening. Between-subjects design used; didn't allow authors to draw conclusions about impact of greening on children's development over time. Data collection limited to one day a year at each school over three |
| | Attention restoration | Digit Letter Substitution Test (DLST) | | At second follow up, significant interaction between time and condition for the improvement in DLST after recess ($p<0.05$) | | | Gender and grade did not moderate effects. | Some support for positive impact on children's attention restoration during recess, after schoolyard has been greened for longer period. | |
| | | Sky Search Task (SST) | | Significant difference in means between intervention and control groups ($p<0.05$) at second follow up.<br>Trend for interaction between time and condition for improvement in SST, $p = 0.08$ | | | | | |
| | Pro-social orientation | Social orientation choice card (SOCC) | | No significant interactions between time and condition at both follow ups, $ps > .25$ | | | Grade moderated impacts of greening on prosocial orientation at first follow up, with a | Some support for positive short-term impact of greening the school on younger, and negative impact on | |

| | | | | | | | | | | |
|---|---|---|---|---|---|---|---|---|---|---|
| | | | | | | | significant interaction between grade 6, condition and time, z = -2.53, p < 0.05. This interaction effect was not present at second follow up. | older children's prosocial orientation. | consecutive years. | |
| | Self-reported social behaviour | Dutch version of SDQ (subscales peer problems and prosocial behaviour) + subscale "social support in friendships" from Dutch social functioning instrument | | Main effects of time were qualified by interactions between time and condition for peer problems only at first follow up (p<0.05), and for social support at both first and second follow up (p<0.001) . No significant interactions between time and condition for self-reported prosocial behaviour, ps > 0.2. | | | At first follow up significant interactions with condition and time for grade 5, z = 2.55, p < 0.05 and for grade 6, z = 2.88, p < 0.05. | Positive impact of greening on some aspects of social functioning, in particular for social support and self-reported peer problems, but not for self-reported prosocial behaviour. | | |
| Waliczek (2001) | Interpersonal relationships | Self-Report of Personality Scale from the Behaviour Assessment | Paired t-tests Independent t-tests ANOVA | No statistically significant differences found in comparisons of differences between pre and post-tests of children in gardening group. | | | Gender did not have a significant effect on outcomes in interperson | | Consistency of intervention, frequency and duration of each | Weak |

| | | | | | | | | | | |
|---|---|---|---|---|---|---|---|---|---|---|
| | | System for Children (BASC) (statements concerning interpersonal relationships and attitude towards school) | | | | | al relationships, but grade did have an effect ( F = 4.186, df = 6, p = 0.000). Seventh grade students (ages 12-13) had the most positive interpersonal relationship scores of all grades. | | episode not reported. Single subjective (self-reporting) measure used; data cannot be corroborated. Differences in changes in variables from pre-test to post-test between intervention and control group not significant. | |
| Wallner (2018) | Wellbeing | Self-condition scale by Nitsch (6 dimensions) | Linear contrasts Kolmogorov-Smirnov tests Mauchly's test Box M tests ANOVA | For all dimensions of the measurement scale, and for all green exposure types (small park, large park, forest), there were significant differences (p<0.05) between time points. The condition of the pupils showed a trend to improve shortly after arrival in the respective green space and mostly reached a significant | | | | No significant differences in wellbeing scores for the initial measurement times, but towards the end of the measurement period only the forest provided/evoked sustained effects in some | Results were confounded by using a self-rating method rather than observation or physiological measures. All sites were green spaces; there was no comparison with built | Weak |

| | | | | | | | | | |
|---|---|---|---|---|---|---|---|---|---|
| | | | | improvement after a longer stay. Statistically significant differences (and in one case a trend towards significance) between the three green space types were found for the dimensions recuperation, tension/relaxation, state of mood, readiness for action, and readiness for exertion. For the forest type, the decrease of wellbeing after return into the classroom was significantly less expressed than after the stay in the small or large urban park (readiness for action: $p < 0.001$; readiness for exertion: $p = 0.027$; state of mood: $p < 0.001$; tension/relaxation: $p < 0.001$) and a trend was found for recuperation ($p = 0.089$). | | | | wellbeing variables (especially in the domains readiness for exertion, mood, and tension/relaxation. | environment or blue space. As all groups had a study break it is impossible to separate effects of break itself and effect of green spaces. Generalisability limited as sample consisted of students who had completed compulsory education. |
| | Attention | d2-R test | | Concentration performance values | Increase after | | | Highest increase of | |

| | | | | | | | | | | |
|---|---|---|---|---|---|---|---|---|---|---|
| | | | | were significantly higher after pupils' stay in green spaces for all sites (p<0.001) | returning from small urban park was 7.5 (SD 9.7); after the large urban park it was 15.5 (SD 11.7) and after the forest it was 5.3 (SD 11.0). | | | performance found for the larger park type, with this increase also being significantly higher (p=0.008) than the increase after stays in the other green spaces. | | |
| Whitburn (2023) | Life satisfaction | Students' Life Satisfaction Scale | Mixed-design ANOVAs | F (1,255) = 2.79, p = 0.10 | | | | No significant change | Authors comments that intervention duration potentially too short to have an impact. Data is from self-report only. Retention rate was only 79.3% for the post-survey at 4 weeks post-intervention. Lower-than ideal statistical | Strong |

| | | | | | | | | | | |
|---|---|---|---|---|---|---|---|---|---|---|
| | | | | | | | | | power (70%) due to drop-outs. Control group was smaller and selected from fewer schools. | |
| | Vitality | Vitality scale adapted from Ryan and Frederick | | $F(1,255) = 0.26$, $p = 0.61$ | | | | No significant change | | |
| Wood (2014) | Self-esteem | Rosenberg Self-esteem Scale), 10 items, modified for age group | Independent t-tests Mixed ANOVA | No significant effect for change in self-esteem due to the environment ($p > 0.05$; $\eta_p^2 = 0.000$) | | | No significant interaction effect for environment and gender ($p > 0.05$); $\eta_p^2 = 0.000$. | Playing in a natural environment not found to have benefit for children's self esteem | Playing within a natural environment may have been insufficient to promote engagement and hence children may have derived limited benefit. Rosenberg self-esteem scale open to ceiling and floor effects. The scale is predominantly used as a trait scale and may have had | Moderate |

| | | | | | | | | | |
|---|---|---|---|---|---|---|---|---|---|
| | | | | | | | | | limited sensitivity in this context. |
