## Supplemental file E Quality Appraisal for "Impacts of green space interventions in educational settings on children and young people’s mental wellbeing: a systematic review"

**Quality appraisal of included quantitative studies**

| First author (year) | EPHPP Criteria for quantitative studies | | | | | | |
|---|---|---|---|---|---|---|---|
| | Selection bias | Study design | Confounders | Blinding | Data collection | Withdrawals and dropouts | **Overall rating** |
| Amicone (2018) | Moderate | Moderate | Strong | Weak | Moderate | Strong | **Moderate** |
| Anabirtate (2021) | Strong | Moderate | Strong | Moderate | Strong | Weak | **Moderate** |
| Barrable (2021) | Weak | Moderate | Weak | Weak | Moderate | Moderate | **Weak** |
| Barton (2015) | Weak | Moderate | Moderate | Weak | Strong | Weak | **Weak** |
| Bates (2018) | Weak | Weak | Weak | Weak | Weak | Weak | **Weak** |
| Block (2012) | Moderate | Moderate | Weak | Weak | Strong | Moderate | **Weak** |
| Brussoni (2017) | Moderate | Moderate | Strong | Weak | Strong | Strong | **Moderate** |
| Carrus (2015) | Moderate | Weak | Weak | Moderate | Moderate | Weak | **Weak** |
| Dopko (2019) | Weak | Moderate | Moderate | Weak | Weak | Weak | **Weak** |
| Ernst (2019)(1) | Weak | Moderate | Weak | Weak | Strong | Weak | **Weak** |
| Ernst (2019(2)) | Weak | Moderate | Weak | Weak | Strong | Weak | **Weak** |
| Harvey (2020) | Weak | Weak | Weak | Weak | Strong | Strong | **Weak** |
| Kelz (2015) | Weak | Moderate | Weak | Moderate | Moderate | Moderate | **Weak** |
| Largo-Wight (2018) | Moderate | Moderate | Moderate | Moderate | Weak | Strong | **Moderate** |
| Mason (2022) | Weak | Weak | Moderate | Moderate | Strong | Weak | **Weak** |
| Montgomery (2022) | Weak | Weak | Weak | Moderate | Strong | Weak | **Weak** |
| Moula (2023) | Moderate | Moderate | Weak | Weak | Strong | Weak | **Weak** |
| Mueller (2017) | Weak | Moderate | Weak | Moderate | Moderate | Weak | **Weak** |
| Mygind (2009) | Weak | Weak | Weak | Weak | Weak | Moderate | **Weak** |
| Pirchio (2021) | Moderate | Moderate | Weak | Weak | Weak | Weak | **Weak** |
| Pollin (2021) | Weak | Weak | Weak | Weak | Weak | Weak | **Weak** |
| Raney (2019) | Weak | Moderate | Strong | Moderate | Strong | Moderate | **Moderate** |

| | | | | | | | |
|---|---|---|---|---|---|---|---|
| Raney (2021) | Moderate | Moderate | Strong | Moderate | Strong | Weak | **Moderate** |
| Reed (2013) | Moderate | Moderate | Moderate | Weak | Strong | Strong | **Moderate** |
| Roberts (2017) | Moderate | Moderate | Moderate | Weak | Strong | Strong | **Moderate** |
| Roe (2011) | Weak | Weak | Moderate | Weak | Strong | Weak | **Weak** |
| Scogin (2023) | Moderate | Moderate | Weak | Weak | Strong | Strong | **Weak** |
| Sheldrake (2019) | Moderate | Weak | Weak | Weak | Weak | Weak | **Weak** |
| Sprague (2020) | Moderate | Weak | Weak | Weak | Strong | Strong | **Weak** |
| Sprague (2021) | Weak | Moderate | Moderate | Moderate | Strong | Weak | **Weak** |
| Taylor (2020) | Moderate | Moderate | Strong | Moderate | Strong | Strong | **Strong** |
| Van den Berg (2024) | Moderate | Weak | Strong | Strong | Strong | Moderate | **Moderate** |
| Van Dijk-Wesselius (2018) | Moderate | Moderate | Moderate | Weak | Strong | Weak | **Weak** |
| Waliczek (2001) | Weak | Weak | Weak | Weak | Weak | Weak | **Weak** |
| Wallner (2018) | Weak | Moderate | Moderate | Weak | Weak | Strong | **Weak** |
| Whitburn (2023) | Moderate | Moderate | Strong | Moderate | Strong | Strong | **Strong** |
| Wood (2014) | Weak | Moderate | Moderate | Moderate | Moderate | Moderate | **Moderate** |
